# Childhood emotional symptom trajectories in three generationally and socio-ethnically distinct UK birth cohorts

**DOI:** 10.64898/2026.06.24.26356453

**Authors:** Sophie J Fairweather, Alex S F Kwong, Emre Deniz, Gemma Hammerton, Golam M Khandaker, Hannah J Jones

**Author notes:** Joint senior authors (Golam M Khandaker, Hannah J Jones). **Corresponding author** Sophie J Fairweather; Population Health Sciences, Bristol Medical School, Augustine’s Courtyard, Orchard Lane, Bristol BS1 5DS, UK.

## Abstract

**Background:** Depression/anxiety symptoms emerge early in life. We examined emotional symptom trajectories, from early childhood, in three UK birth-cohorts spanning successive generations and diverse socio-ethnic contexts.

**Methods:** We used data from the Avon Longitudinal Study of Parents and Children (ALSPAC), Millennium Cohort Study (MCS) and Born in Bradford (BiB) to identify group-based emotional symptom trajectories from 3-14y (N=11,025; 15,446; 6,711). Participants were born three decades apart (ALSPAC: 1990-2, MCS: 2000-2, BiB: 2007-10). We characterised groups by: sex, ethnicity, maternal depression/anxiety and IMD quintile. In ALSPAC we modelled associations between trajectories and depression/anxiety diagnoses at 24y and 30y.

**Results:** Participants from ethnic minority backgrounds comprised 4% of ALSPAC, 17% of MCS and 66% of BiB. In all cohorts 49% were female. Each cohort had low-, mid- and high-level symptom trajectories. High-level trajectories comprised 6-7% of children. In younger cohorts (MCS and BiB), the high-level symptom trajectory started high and persisted while in the older ALSPAC cohort it started low and increased. In MCS and BiB, low-to-medium-level symptoms increased over time, but remained stable in ALSPAC. Female sex and maternal depression/anxiety were associated with increased odds of high-level trajectories (all cohorts). Lower area-level deprivation and belonging to the local ethnic majority group were protective. Mid- and high-level symptom trajectories had higher odds of depression/anxiety diagnoses in early adulthood in ALSPAC.

**Conclusions:** Childhood emotional symptom trajectory-groups are broadly similar across generations and diverse social contexts. However, children born after 2000, in more diverse contexts, may experience persistent symptoms from a younger age.

## Introduction

Depression and anxiety symptoms often emerge early in life. Approximately half of depression cases emerge before age 30 and nearly 40% of anxiety cases emerge before age 14 (Solmi et al., 2022). It is widely reported that prevalence of depression and anxiety in the population is increasing, especially in young people (Hua et al., 2024; Liu et al., 2024). In the UK, an increase in primary care reported anxiety and depression has been observed in children as young as six (Cybulski et al., 2021). However, much of the existing research into developmental trajectories of depression and anxiety in young people has focused on adolescence and early adulthood (Shore et al., 2018, Portogallo et al., 2024).

Evidence from population growth models suggest anxiety/depression are manifesting earlier, lasting longer, and increasing at a faster rate in children born in 2000s vs those born in the 1990s, in the UK (Armitage et al., 2023). However, studies investigating which sub-groups contribute to overall population level growth are lacking. For example, we do not know whether the prevalence of children with persistent, high-level emotional symptoms has changed across generations.

Diverse, contemporary cohorts, using the same assessment method longitudinally are available but have so far been underutilised in comparative cross-cohort trajectory studies. Especially cohorts in which participants are currently of adolescent age (born around 2010-12). Adolescents of this age grew up amidst important social and economic pressures that have been suggested as possible determinants of mental health trends in young people, such as prevalent smart phone use, rising inequalities, cuts to youth services, educational forms, and the COVID-19 pandemic (McGorry et al., 2025).

Studying long-term trends of early pre-clinical emotional symptoms is critical because clinical anxiety and depression often first manifest as emotional difficulties in childhood. Capturing early onset and longitudinal trajectories can help inform strategies for prevention, intervention, and resource allocation involving health, education, social care and other agencies to improve long-term population-level outcomes. Specifically, identifying groups of children with early-onset, persistent emotional problems in childhood through to adolescence. Prior meta-analysis of longitudinal depressive symptom trajectories in childhood/adolescence shows that persistent emotional symptoms in early life predict later diagnoses of major depressive disorder (MDD) (Portogallo et al., 2024) alongside a range of poor social and physical health outcomes, for example NEET (“Not in education, employment or training”) status (Morales-Muñoz et al., 2023; Rahmani & Groot, 2023).

In the current study we aimed to: a) identify a high/persistent emotional symptom trajectory group spanning early-childhood to early-adolescence in three generationally, ethnically, and socioeconomically diverse UK cohorts; b) compare group size/proportions, symptom severity over time and onset across cohorts; and c) characterise trajectory groups by sociodemographic factors, and their association with mental illness diagnosis in early-adulthood.

## Methods

### Study design and participants

We used data from three successive, prospective UK birth cohorts born between 1990 and 2010: The Avon Longitudinal Study of Parents and Children (ALSPAC), Millenium Cohort Study (MCS) and Born in Bradford (BiB). Baseline samples in our primary analyses comprised 11,025 children from ALSPAC (born 1990-92), 15,446 children from MCS (born 2000-02) and 6711 children from BiB (born 2007-10) with ≥one parent/teacher reported Strengths and Difficulties Questionnaire (SDQ) measure between age 3-14y. In all cohorts we removed siblings keeping only one childhood per nuclear family at random.

Ethical approval for each cohort was obtained from the relevant Local Research Ethics Committee and informed consent was obtained from participants, full details for each cohort can be found in the Supplementary Materials.

**ALSPAC:** enrolled pregnant women living in Avon, UK with expected delivery dates between 1st April 1991 and 31st December 1992. The initial number of pregnancies 14,541 and 13,988 children were alive at age 1y. When children were approximately 7y, the initial sample was bolstered with further eligible cases. The total sample age 7y is 15,447. Of these 14,901 children were alive at age 1y (Boyd et al., 2013; Fraser et al., 2012; Northstone et al., 2019, 2025). The study website contains details of available data through a searchable data dictionary and variable search tool: http://www.bristol.ac.uk/alspac/researchers/our-data/.

**MCS:** enrolled 18,827 babies (age 9 months) from 18,552 families born in England or Wales between 1^st^ September 2000 and 31^st^ August 2001, and Scotland or Northern Ireland between 23^rd^ November 2000 and 11^th^ January 2002. Children from disadvantaged areas and ethnic minority backgrounds were over-sampled to ensure the sample was nationally representative. MCS data are available via the UK Data Service [SN 2000031] (UK Data Service, 2024).

**BiB**: is a multi-ethnic birth cohort study situated within the City of Bradford (McEachan et al., 2024). 12,453 pregnant women were recruited between 2007-2011 during routine glucose testing appointments, resulting in an initial cohort of 13,776 children. Several sub-cohorts have been enrolled which include original participants and additional non-BiB recruits. In this study we have used data from two classroom-based sub-cohorts: Starting School (4-5y) and Primary School Years (7-8y), and two follow-up studies: Growing Up (7-9y) and Age of Wonder which is a seven-year project that commenced in academic year 2021-2022, following BiB and non-BiB children through their teenage years. We used original BiB participants only.

### Measures

#### Emotional symptom measure

Emotional symptoms were measured using the emotional sub-scale of the Strengths and Difficulties Questionnaire (SDQ-E; range 0-10). SDQ-E is a widely used measure of child mental health. Questions include worries/fears, depressed mood and somatic complaints. SDQ-E has good concurrent and discriminant validity (Goodman, 2001; Lundh et al., 2008; Muris et al., 2003), and performs well in validation studies for detecting anxiety and depression cases across development (Armitage et al., 2023).

SDQ-E was measured at multiple timepoints between age 3-17y. To harmonize timepoints across cohorts we grouped the SDQ-E observations into age five age bands: 3-5, 6-8, 9-11, 12-14, and 15-17 years. In the case of multiple measures for an individual within an age band the first measure was taken. Our primary models used data from 3-14y in all cohorts as data were sparse at 15-17y in BiB.

In ALSPAC and MCS, SDQ-E was parent-reported at all timepoints. In BiB SDQ-E was teacher reported at two waves, parent reported in one wave and self-reported in Age of Wonder. Detailed information regarding measures and reporter type by age bands can be seen in the Supplementary Materials (Tables S1a, 1b), along with sensitivity analyses addressing possible measurement differences between reporter types.

### Sociodemographic variables and maternal depression/anxiety

Sociodemographic measures include child sex at birth/early infancy, maternal depression and/or anxiety, ethnic group, and 2010 Index of Multiple Deprivation (IMD quintiles). The IMD ranks areas from most to least deprived across seven domains (income, employment, education, health, crime, living environment and housing). Due to data sparsity we dichotomized ethnicity as white/ethnic minority groups. White includes: English, Welsh, Scottish, Northern Irish or British; Irish; any other White background. Ethnic minority groups include: Mixed/multiple, Asian or Asian British, Black, Black British, Caribbean or African, Arab, Other ethnic groups. Data were reported by mothers in questionnaires.

### Mental health diagnoses at age 24 and 30 years in ALSPAC

In ALSPAC we examined associations between trajectory groups and binary ICD-10 diagnosis of depression and presence of generalized anxiety disorder (GAD) at ages 24 and 30y. Cases were derived from the self-administered computerized Clinical Interview Schedule-Revised (CIS-R) (Fairweather et al., 2026).

### Statistical analyses

Data was cleaned in STATA v19.0 (StataCorp, 2025). We used Mplus (v8.11) (Muthén, B. O., 2017) to model trajectories.

#### Trajectory modelling

Our objective was to identify clinically meaningful and interpretable trajectory groups in each cohort, with primary interest in identifying a high/persistent symptom trajectory. Inclusion of BiB as a contemporary, ethnically diverse cohort was key. Emotional symptom trajectories have not been modelled in this cohort before. We used Latent Class Growth Analysis (LCGA) to derive data-driven latent classes from clusters of individuals with similar patterns of change in SDQ-E over time. Parsimony was a priority given the smaller sample in BiB. Alternative modelling approaches were considered; see Supplementary Materials. Supplementary figure S3b shows that a linear model fits the observed means well. We adhered to the Guidelines for Reporting on Latent Trajectory Studies (van de Schoot et al., 2017) (GRoLTS; checklist in Supplementary Materials).

Class posterior probabilities were used to assign individuals to trajectories. We preserved the relative spacing between timepoints and retained a zero anchor. Identical methods were used across cohorts to ensure we limited variation from sources other than the data.

All models were estimated using maximum likelihood which is robust to non-normality. We used 1000 random starts, 200 final iterations. The best log-likelihood was replicated at least 20 times in all models. We considered class solutions from one to six classes. Final models were selected based on clinical interpretability/meaning of groups, comparison of model fit statistics, and sample size of the smallest class. We aimed to avoid very small classes (<5% of the overall sample). Fit statistics included: class average posterior probabilities, Akaike Information Criterion (AIC), Bayesian Information Criterion (BIC), sample-adjusted-BIC, entropy and Lo-Mendell-Rubin likelihood-ratio-test (LMR-LRT).

Participants contributed to a model if they had at least one datapoint. Full information maximum likelihood (FIML) was used to fit the model, assuming data are missing at random. See Supplementary Materials for full details of missing data handling and distributions (Supplementary Tables S2a-2c).

#### Cross cohort comparisons

After estimating trajectories in each cohort individually, we combined data from the three cohorts and formally tested for cross-cohort differences using a categorical variable to define cohort membership. We used the original, cohort-specific derived slope and intercept estimates as starting values for each trajectory group in a combined model and allowed class proportions to differ between cohorts. We performed pairwise Wald tests to test for evidence of differences between slopes and intercepts of the corresponding classes across cohorts (e.g., ALSPAC high vs MCS high). P-values were adjusted for multiple testing using the Bonferroni method (Dunn, 2012).

#### Subgroup characterisation

We used the bias-adjusted-three-step method (Asparouhouv & Muthen, 2014) to characterise latent trajectories by sex, ethnicity, IMD, and maternal depression/anxiety, and to study associations between trajectory class and depression/anxiety diagnoses at ages 24 and 30 in ALSPAC. This method accounts for classification uncertainty while preventing covariates from redefining classes or artificially inflating entropy. Samples for univariable regression models included participants with complete data on the characterising factor and the latent class (hence sample size varied between models).

#### Sensitivity analyses

We ran supplementary analyses in MCS and ALSPAC using data from age 3-17y (Supplementary Materials, Figure S2). In all cohorts we conducted sensitivity analyses restricting the sample to participants with ≥2 repeated measures (Supplementary Materials, Figure S3a).

Given the multiple reporters in BiB (school, parent and self; Table S1b), we conducted sensitivity analyses to test whether original BiB trajectory groupings were an artefact of reporter type by standardising SDQ-E scores across reporter types in two ways.

First, at each timepoint we calculated mean differences between the observed BiB SDQ-E scores (teacher- or self-reported) and published parent-reported “norms” at ages corresponding to the timepoints in our study (Table S7). These “norms” are based on a large national survey of child and adolescent mental health carried out by the Office for National Statistics (Meltzer et al., 2000).

Second, where there were both parent/teacher or parent/self-reported SDQ-E scores at a single timepoint in BiB, we calculated mean differences between the teacher/self-reports compared to the parent reported means at the same timepoint (Table S8).

Based on these mean differences, we calculated two sets of SDQ-E scores standardised to parent-reported levels (standardised to parent-reported “norms” and standardised to observed BiB parent-reports) by adjusting the non-parent reported scores up or down accordingly. We then re-ran the three-class LCGA models using each set of adjusted SDQ-E scores, and compared the slopes, intercepts and 95%CIs of the classes to those from the original three-class LCGA model (estimated using observed SDQ-E scores)(Supplementary Figure, S1).

## Results

### Sample characteristics

Baseline samples in our primary analyses comprised 11,025 children from ALSPAC (born 1990-92), 15,446 children from MCS (born 2000-02) and 6711 children from BiB (born 2007-10) with ≥1 parent/teacher reported Strengths and Difficulties Questionnaire (SDQ) measure between age 3-14y.

In all cohorts 49% of the baseline sample were female (Table 1). ALSPAC has very few ethnic minority participants (4%) compared to MCS (17%) and BiB (66%). Over two-thirds of BiB participants were in the most deprived IMD quintile, compared to 35% of MCS participants and 14% of ALSPAC participants. Roughly 25% of mothers had depression/anxiety in both ALSPAC and MCS. Nearly half of mothers had depression/anxiety in BiB.

**Table 1.** Baseline characteristics.

|  | ALSPAC | MCS | BiB |
|---|---|---|---|
| Baseline sample | 11,025 | 15,446 | 6,711 |
| % Female | 5373/11004 (49%) | 7252/14818 (49%) | 3278/6711 (49%) |
| Ethnic group |  |  |  |
| Ethnic minority groups | 410/9943 (4%) | 2496/14783 (17%) | 4422/6710 (66%) |
| IMD quintile |  |  |  |
| Q1 (most deprived) | 854 / 6229 (14%) | 3194 / 9146 (35%) | 3822 / 5618 (68%) |
| Q2 | 1138 / 6229 (18%) | 1978 / 9146 (22%) | 985 / 5618 (18%) |
| Q3 | 1117 / 6229 (18%) | 1567 / 9146 (17%) | 605 / 5618 (11%) |
| Q4 | 1446 / 6229 (23%) | 1260 / 9146 (14%) | 137 / 5618 (2%) |
| Q5 (least deprived) | 1674 / 6229 (27%) | 1147 / 9146 (13%) | 69 / 5618 (1%) |
| Maternal depression or anxiety |  |  |  |
| Yes | 2845 / 11025 (26%) | 3623 / 14791 (24%) | 2054/4737 (43%) |
| Mean SDQ-E score (SD) |  |  |  |
| 3-5 years | 9166<br>1.45 ± 1.52 | 14,619<br>1·41 ± 1·61 | 2167<br>1·05 ± 1·65 |
| 6-8 years | 8995<br>1.66 ± 1.82 | 12,055<br>1·54 ± 1·78 | 1372<br>1·96 ± 2·10 |
| 9-11 years | 8606<br>1.51 ± 1.76 | 11,417<br>1·87 ± 2·00 | 3866<br>2.21 ± 2.17 |
| 12-14 years | 7039<br>1.44 ± 1.72 | 10,021<br>2.05 ± 2.14 | 1223<br>3.70 ± 2.74 |
| Trajectory membership |  |  |  |
| Low-level symptom trajectory | 72% | 73% | 75% |
| Mid-level symptom trajectory | 22% | 21% | 18% |
| High-level symptom trajectory | 6% | 6% | 7% |
| Prevalence of mental health diagnoses |  |  |  |
| Depression<br>(age 24) | 369/3617 (10%) | NA | NA |
| GAD<br>(age 24) | 352/3608 (10%) | NA | NA |
| Depression<br>(age 30) | 334/3365 (10%) | NA | NA |
| GAD<br>(age 30) | 442/3354 (13%) | NA | NA |

Footnotes: ALSPAC=Avon Longitudinal Study of Parents and Children, MCS=Millenium Cohort Study, BiB=Born in Bradford, IMD=index of multiple deprivation, Q=quintile, SDQ-E=emotional subscale of the Strengths and Difficulties Questionnaire, SD=standard deviation, GAD=generalized anxiety disorder, NA=not applicable.

### Selection of emotional symptom trajectory groups

A three-class solution was selected. A high-level symptom group was identified in all cohorts by a three-class model. Group sizes became compromised (even in the cohort with the largest sample) when additional classes were added (the four class model in MCS identified a group size <4%; Table S4). In the three-class model, average posterior probabilities exceeded 0.80 for all classes. Entropy was moderate (0.79 in MCS and ALSPAC; 0.64 in BiB). Entropy was only moderately improved in one cohort by identification of a fourth class. Information criteria (AIC, BIC, SABIC) improved with additional classes, although the magnitude of improvement diminished with further classes and class patterns were not clinically relevant to our research aim (Supplementary Material, Tables S3-5).

### Similarities and differences between cohorts

Despite generational and sociodemographic differences between cohorts, we observed a high-level symptom trajectory and similar proportions of trajectory group membership in all cohorts. Between 72-75% of each cohort belonged to a low-level symptom trajectory (scores <2), 18-22% of each cohort belonged to a mid-level symptom trajectory (scores 3-4), and approximately 6-7% belonged to a high-level symptom trajectory (scores ≥5) (Figure 1, Table 1).

**Figure 1.**
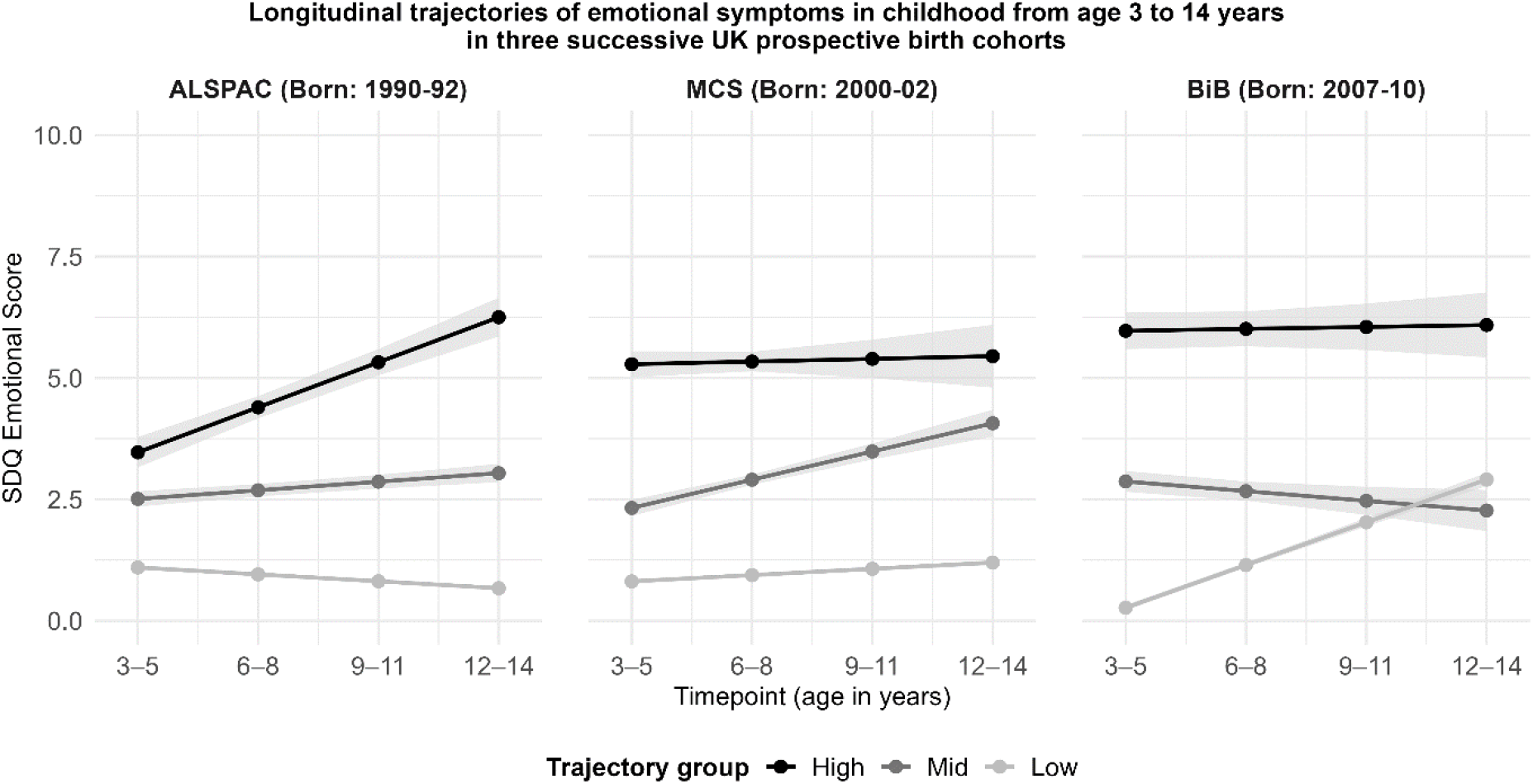
Longitudinal trajectories of emotional symptoms in childhood from age 3 to 14 years in three successive UK prospective birth cohorts.

Two key differences were observed between cohorts. In the high-level symptom trajectory, symptom level started high and remained persistent across time in the younger cohorts born after 2000 (MCS and BiB) whereas symptom level in this group started low and steeply increased in the oldest cohort (born early 1990s; ALSPAC). In the younger cohorts, born after 2000 (MCS and BiB), either the mid- or low-level symptom trajectories increased over time, unlike ALSPAC where these groups were stable.

In MCS (born 2000) the mid-level trajectory increased and, by age 12-14 years symptom level nearly reached that of the high-level trajectory group. In BiB (the youngest cohort; born 2007-10) the low-level trajectory increased and, over time, overlapped with the mid-level trajectory. However, the high-level trajectory group shows clear separation from the mid- and low-level trajectories (Figure 1, Table S15).

Results of pairwise Wald tests comparing high, mid, and low classes across cohorts support our descriptive findings and trajectory group characterisation (Supplementary Table S6a). For all but one of the comparisons there was strong evidence that slopes and intercepts differed between cohorts. The exception being slopes of the high classes in MCS and BiB where there was little statistical evidence that these differed; i.e., both are persistently high across time (p=0.95; Figure 1). In contrast, there was strong statistical evidence that the slope of the high-class in ALSPAC differed to both MCS, and BiB high-class slopes (both comparisons p<0.001).

### Sociodemographic correlates of trajectory groups

In all cohorts, females and children of mothers with depression/anxiety had increased odds of being in high-level or mid-level symptom trajectory groups (Figure 2, Tables S14-15).

**Figure 2.**
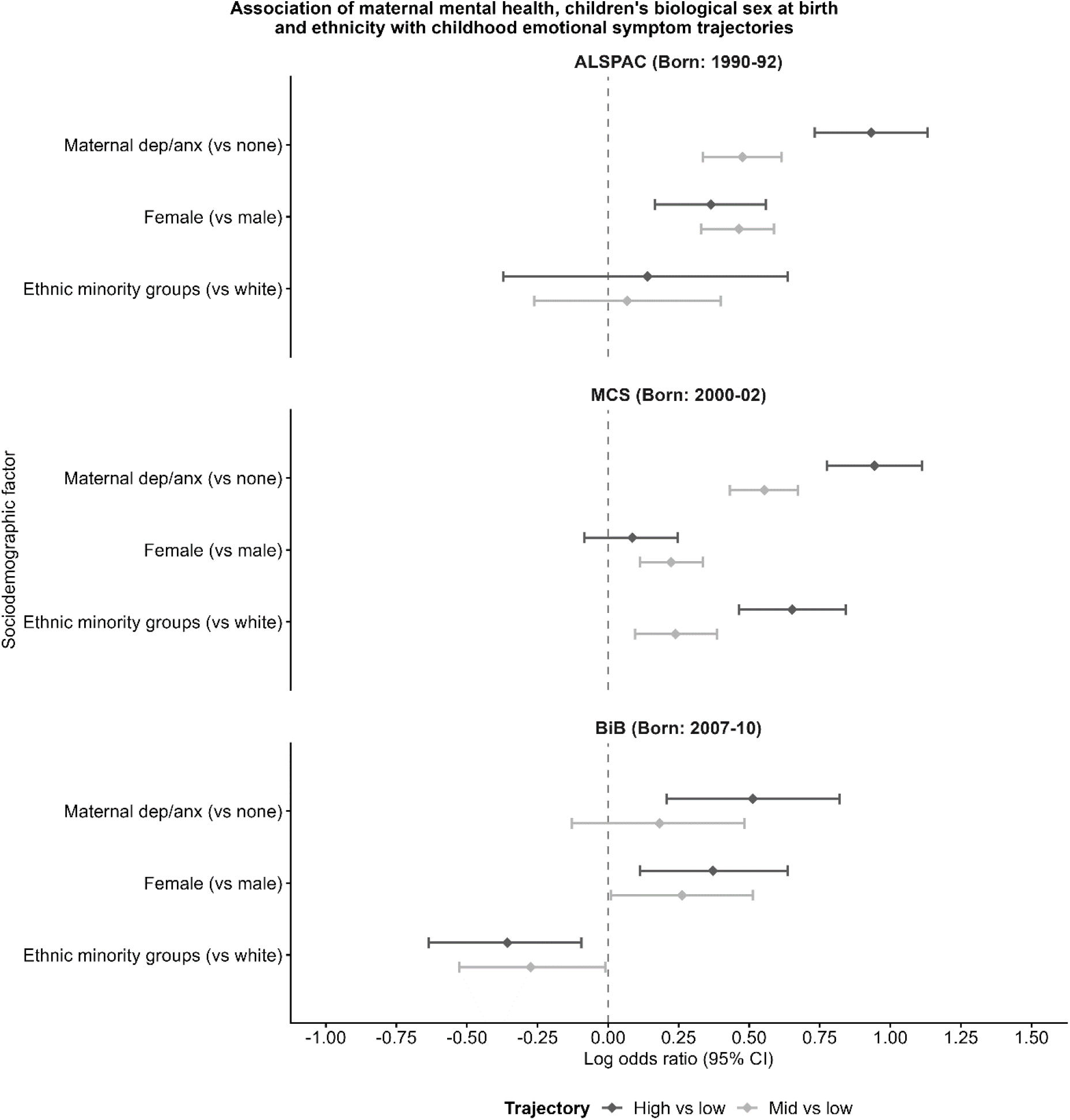
Association of maternal mental health, children’s sex at birth and ethnicity with childhood emotional symptom trajectories.

Ethnicity showed opposing associations with emotional symptom trajectories between cohorts. In MCS, odds of high-level emotional symptoms (vs low-level) were nearly double in the ethnic minority participants vs. white participants (majority group). In BiB this was reversed; belonging to an ethnic minority group (the local majority group in Bradford) was protective against both mid- and high-level symptom trajectory membership (Figure 2, Tables S14-15).

Higher area-level deprivation was strongly linked to high-level emotional symptoms across childhood. In MCS we observed a dose–response pattern. A similar pattern was seen across symptom trajectories: lower deprivation was more protective for children with high-level symptoms compared to low-level symptoms. In ALSPAC, this pattern was mainly seen for the high-level vs low-level symptom group comparison, but not in the mid-level comparison, likely because this cohort is less socioeconomically diverse. In BiB, there was little evidence overall of an association due to small samples and large CIs (Figure 3, Tables S14-15).

**Figure 3.**
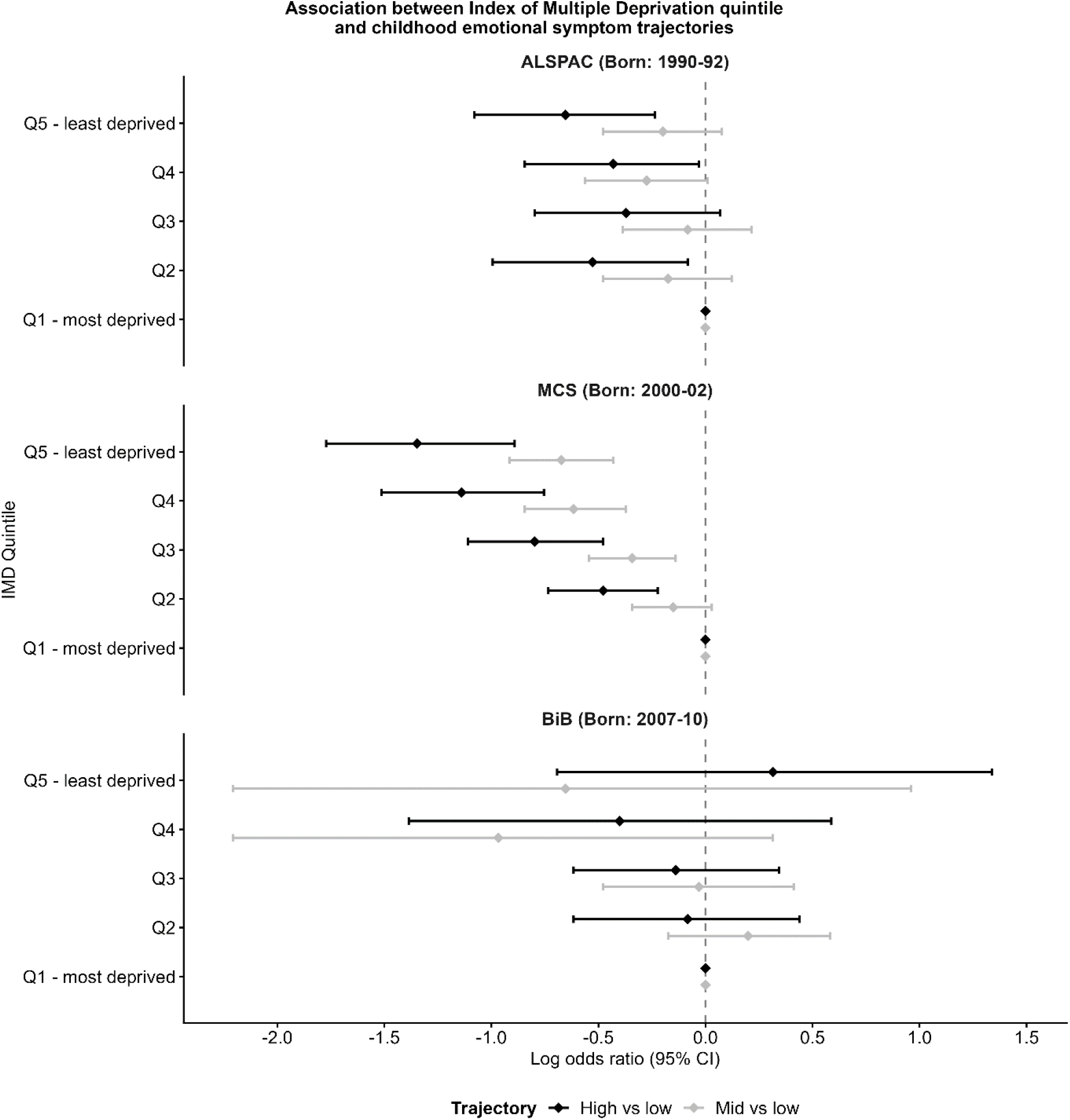
Association between Index of Multiple Deprivation quintile and childhood emotional symptom trajectories.

### Association with depression and anxiety in adulthood

High- and mid-level symptom trajectories were associated with increased odds of depression and GAD at age 24 in ALSPAC (Table 2). Compared to the low-level symptom trajectory, the effect size for the high-level symptom trajectory was larger than that for the mid-level (vs. low-level) symptom trajectory. Similarly, the mid-level symptom trajectory was associated with higher odds of GAD and depression at age 30. The high-level symptom trajectory was associated with increased odds of GAD, however, there was no strong evidence of association with depression at age 30.

**Table 2.** Associations of childhood emotional symptom trajectories with ICD-10 depression or generalized anxiety disorder (GAD) in the ALSPAC cohort at ages 24 and 30.

|  |  | Analysis sample size | Symptom trajectory, Odds ratio (95% CI) |  |  |
|---|---|---|---|---|---|
|  |  |  | Low | Mid | High |
| 24y | Depression | 3617 | 1.00 (Reference) | 1.50 (1.07-2.10) | 2.74 (1.70-4.42) |
|  | GAD |  |  | 1.79 (1.28-2.49) | 2.33 (1.38-3.93) |
| 30y | Depression | 3365 | 1.00 (Reference) | 1.52 (1.08-2.14) | 0.82 (0.39-1 .72) |
|  | GAD |  |  | 1.84 (1.35-2.51) | 2.61 (1.68-4.08) |

### Results of sensitivity analyses

In sensitivity analyses ALSPAC and MCS trajectory groups remained broadly unchanged when a further datapoint was added at age 15-17 (Supplementary Material, Figure S2) and when primary analyses were restricted to participants with ≥2 repeated measures (Supplementary Material, Figure S3a). In BiB, the high-level trajectory changed when the sample was restricted to participants with ≥2 repeated measures. However the remaining sample was very small at the final timepoint making the estimate difficult to interpret (Supplementary Material, Figure S3a). In BiB, trajectory group patterns were broadly unchanged in sensitivity analyses after adjusting SDQ-E scores to account for differences in reporter type across timepoints (Supplementary Material; Figure S1).

## Discussion

We identified and characterised group-based early-childhood trajectories of emotional symptoms in three ethnically and socioeconomically diverse successive UK population-based cohorts, born two decades apart. Despite generational, ethnic and socioeconomic differences, we identified a high-level symptom group of a similar prevalence (6-7%) in all three cohorts.

Two key differences emerged. In the high-level symptom trajectory, symptom level in childhood starts high and persists across time in the younger cohorts whereas in the oldest cohort (born in the early 1990s) symptoms start low and steeply increase. In the younger cohorts, born after 2000, mid- or low-level symptom trajectories show an increase in symptom level, and become closer to mid/high groups over time, unlike the oldest cohort (born early 1990s) where these groups are stable. Considering both high-level and increasing symptom trajectories together, a larger proportion of children in the younger, more diverse cohorts belonged to these groups.

These findings support existing evidence that in recent decades emotional symptoms are increasing among children from an early age. Particularly in groups that experience relatively low-level symptoms, while symptom severity in group of children experiencing high-level of symptoms remains stable. Therefore, the overall picture is consistent with an increase in low-to-mid level symptoms in young children in recent decades, aligning with wider data trends in the UK showing a sharp rise in the incidence of depression and anxiety in young people from as young as six years old since 2012 (Cybulski et al., 2021).

Across all cohorts, female sex and maternal depression/anxiety were associated with higher odds of persistent or increasing symptom trajectories, whereas higher SES and belonging to the local ethnic majority were protective factors.

Prevalence of persistent-high symptoms aligns with broader trajectory literature despite differences in age, geography, and psychiatric measure. Group-based trajectory modelling studies identified in a large systematic review (Shore et al., 2018) mostly studied US populations with trajectories starting later (in early/mid adolescence). Studies typically identify similar patterns with <10% prevalence in the persistent group and are mostly of depression trajectories which might explain the slightly higher prevalence in persistence as depression onsets later than anxiety (Solmi et al., 2022). Few studies have modelled early group-based emotional symptom trajectories, capturing pre-clinical symptoms measured by screening questionnaires. Two previous studies which have modelled SDQ-E trajectories in ALSPAC from age 4-17 found 6% in the persistent group, consistent with our findings (Dachew et al., 2023; Tseliou et al., 2024). Studies that have *compared* SDQ trajectories across multiple UK cohorts have examined overall population growth rather than investigating group-based trajectories (Armitage et al., 2023; McNicholas et al., 2024). Our findings build upon these, suggesting that that overall population growth may be attributed to symptom increases in children with initially mild to moderate symptoms rather than increased prevalence of children experiencing persistently high emotional symptoms from an early age.

Like our findings, previously identified predictors of trajectories with greater symptom burden include female gender, maternal depression/anxiety, lower income/education and non-white ethnicity (Dachew et al., 2023; Shore et al., 2018; Tseliou et al., 2024). However, research frequently excludes underrepresented populations and ethnic minority groups which are often at highest risk. Our findings build on these by comparing findings in ALSPAC (a largely white, higher SES cohort) with trajectories in two more diverse UK cohorts (MCS and BiB).

In the BiB cohort, belonging to an ethnic minority group was associated with lower odds of persistent high symptoms which contrasts with findings in other cohorts. Bradford, where the BiB cohort is based, has a large community of South Asian heritage. Previous findings in the cohort show South Asian mothers had a lower incidence of common mental health disorders compared to white mothers (Santorelli et al., 2024), our results are consistent with these findings. Protective effects may reflect belonging to the local area ethnic majority, a supportive role of extended family and community networks. However, this observation might also be explained by underreporting of mental health problems in South Asian communities.

Generational differences in social determinants of emotional problems between the cohorts may underpin some of our findings. A review by McGorry et al (McGorry et al., 2025). discusses social trends that might be contributing to rising mental health problems in young people. These include, rising inequalities and economic insecurity, educational reforms, social media/smart phone usage and associated sleep deprivation. Several of these factors disproportionately affect children of lower SES leading to higher emotional problems in this group, which is supported by our findings. In young children effects may be indirect e.g., mediated by parental or home-environment factors like maternal emotional problems (McGorry et al., 2025).

Our findings reinforce the importance of sociodemographic and early life risk factors in predicting mental health trajectories. The two younger cohorts included in this study were more deprived overall which could explain the earlier onset and the higher proportion belonging to persistent/increasing trajectories. Furthermore, rates of maternal depression/anxiety were higher in the youngest cohort and in MCS rates of severe maternal depression/anxiety were comparable to rates of all/any maternal depression/anxiety in ALSPAC. Family psychiatric history and sociodemographic factors were common features in models identified by a recent systematic review of prediction models for anxiety/depression trajectories (Fairweather et al., 2026).

Increased mental health awareness and reduced stigma might partially explain increases in mild sub-clinical emotional symptoms. In the current study we see a fairly stable proportion of young people belonging to a high-risk, persistent group while the proportion in low/mid symptom groups increase. It has been suggested that reduced stigma and increased reporting/awareness of mental health in both young people and the parents of young people is contributing to this trend. Although beneficial, an unintended consequence of reduced stigma and improved awareness of mental health might be wider recognition/reporting of subclinical or transient emotional symptoms that are overinterpreted as clinical anxiety or depression. While this may lead to artefactual increases it has been hypothesised that this might drive genuine increases in new cases and worsening of existing cases by perpetuating behaviours such as avoidance (Foulkes & Andrews, 2023).

In the wider literature there is evidence of increasing severe CMD cases and symptoms (e.g., self-harm) in young people in the UK (Cybulski et al., 2021) as well as milder/moderate self-reported psychological distress. Identifying increasing and persistent emotional trajectories, early, before symptoms escalate is a useful and novel way to differentiate this group from children with mild transient symptoms; as highlighted in a recent *Lancet Psychiatry* Commission on youth mental health (McGorry et al., 2024).

A major strength of our study is replication of findings across three ethnically and socioeconomically diverse populations, including groups frequently underrepresented in mental health research and using a contemporary cohort. We harmonized our modelling approaches across cohorts as far as possible. However, analyses exploring associations between key demographic factors and the persistent trajectory group in the BiB cohort were likely underpowered. Measurement invariance may explain the apparent increase in low-level symptoms, reflecting parent/teacher under-reporting earlier and more accurate self-report later. However, we conducted sensitivity analyses to test this possibility. Broadly trajectory groups were unchanged which was reassuring. The fact that both sex and maternal depression/anxiety associated with high-level vs low-level symptom trajectories in BiB (like the other cohorts) further supports the characterisation of these groups.

High symptom trajectory membership predicted depression/GAD diagnoses in early adulthood (in ALSPAC), strengthening the validity of our trajectory groupings. Anxiety is often under researched compared to depression and our results support further research in this area as we high emotional symptom trajectories in early childhood strongly associates with future GAD diagnoses at age 24 and 30 in ALSPAC. However, we found no evidence that high symptom trajectories predicted depression at age 30. Given the finding at age 24 and strong existing evidence that childhood/adolescent symptoms persist into adulthood (Portogallo et al., 2024), it is likely that this finding reflects selection bias; attrition is a common limitation of longitudinal cohort data and there were substantially fewer depression cases in the high-persistent group at age 30.

In young, contemporary cohorts symptoms are emerging early. Health, education, social care and other services need to be responsive to this emerging trend, which may have long-term implications for young people. Early prediction of children at high-risk of developing persistent symptoms is an opportunity for early intervention and possible improvement of long-term outcomes. This would support better allocation of resources to children with the highest need. However, ethical implications and risks of early screening for emotional symptoms in young people need consideration. Future research also needs to explore individual causal predictors of early-emerging, persistent trajectories, across diverse groups and generations to understand drivers of early onset.

## Conclusion

Our findings show that the prevalence of high-level emotional symptoms in early life remains stable across UK cohorts spanning different generations and diverse ethnic and socioeconomic contexts. However, children in more diverse, contemporary cohorts appear more likely to experience persistently high symptom levels and increases from initially lower symptom trajectories. These findings align with wider data trends in the UK showing a rise in the incidence of depression and anxiety in young people, along with earlier onset in younger generations. Health, education, social care and other services need to be responsive to this emerging trend, which may have important long-term implications for young people.

## Correspondence

Sophie J Fairweather; Population Health Sciences, Bristol Medical School, Augustine’s Courtyard, Orchard Lane, Bristol BS1 5DS, UK

## Acknowledgements

**ALSPAC**: We are extremely grateful to all the families who took part in this study, the midwives for their help in recruiting them, and the whole ALSPAC team, which includes interviewers, computer and laboratory technicians, clerical workers, research scientists, volunteers, managers, receptionists and nurses

**MCS equivalent:** We are grateful for the cooperation of the Millennium Cohort Study families who voluntarily participated in the study. There was no financial compensation for these contributions. We would also like to thank stakeholders from academic, policy maker, and funder communities and colleagues at the Centre for Longitudinal Studies involved in data collection and management of this cohort study.

**Born in Braford:** Born in Bradford is only possible because of the enthusiasm and commitment of the children and parents in BiB. We are grateful to all the participants, health professionals, schools and researchers who have made Born in Bradford happen.

## Ethics statement

**ALSPAC:** Ethical approval was obtained from the ALSPAC Ethics and Law Committee and the Local Research Ethics Committees. Initial cohort approvals:

- Bristol and Weston Health Authority: E1808 Children of the Nineties: Avon Longitudinal Study of Pregnancy and Childhood (ALSPAC) (Date: 28th November 1989)
- Southmead Health Authority: 49/89 Children of the Nineties – "ALSPAC" (Date: 5th April 1990)
- Frenchay Health Authority: 90/8 Children of the Nineties (Date: 28th June 1990)

Informed consent for the use of data collected via questionnaires and clinics was obtained from participants following the recommendations of the ALSPAC Ethics and Law Committee at the time. Participants can contact the study team at any time to retrospectively withdraw consent for their data to be used. Study participation is voluntary and during all data collection sweeps, information was provided on the intended use of data.

**MCS:** Data collection was approved by the UK National Health Service Research Ethics Committee (for MCS1, South West MREC [MREC/01/6/19 – Date 2000/1]; for MCS2 and MCS3, London MREC [MREC/03/2/022 – date 2003/4, 05/MRE02/46 – date 2005/6]; for MCS4, Yorkshire MREC [07/MRE03/32 – date 2007/8]; for MCS5, Yorkshire and The Humber-Leeds East [11/YH/0203 – date 2011/12]; for MCS6, London MREC [13/LO/1786 – date 2015/16]. For MCS7 ethical approval for the pilot surveys and the main stage was obtained by CLS. Ethical approval for the first pilot was obtained on 7th March 2017 from the UCL IOE Research Ethics Committee (ref: REC 884). Approval for the dress rehearsal was obtained on 12th June 2017 from the UCL IOE Research Ethics Committee (ref: REC 951). Ethical approval for the main stage was obtained on 18th October 2017 from the National Research Ethics Service (NRES) Research Ethics Committee (REC) North East – York (REC ref: 17/NE/0341). Written consent was obtained from all parents at each survey.

**BiB:** Ethics approval for the Born in Bradford study was granted by the National Health Service Health Research Authority Yorkshire and the Humber (Bradford Leeds) Research Ethics Committee (reference: 07/H1302/112, date of approval 01/04/2008). Informed consent for data collection and linkage to routine healthcare records was provided by participants at recruitment to the BiB cohort study. Further information on our privacy policy can be found here: https://borninbradford.nhs.uk/privacy-policy/.

## Contributions

SJF, HJJ and GMK designed this study. SJF conducted data analysis, contributed to interpretation of findings, wrote the first draft and carried out manuscript revisions. HJJ contributed to data analysis. HJJ, GH and AK contributed to interpretation of findings, and revised the manuscript. GMK provided supervision for the project including advice on data analysis, interpretation of findings, and revised the manuscript.

## Data availability and data sharing

**ALSPAC:** The ALSPAC study website contains details of all the data that is available through a fully searchable data dictionary and variable search tool: http://www.bristol.ac.uk/alspac/researchers/our-data/. Data used for this submission can be made available on request to the ALSPAC executive committee. The ALSPAC data management plan (available here: http://www.bristol.ac.uk/alspac/researchers/data-access/) describes in detail the policy regarding data sharing, which is through a system of managed open access.

The steps below highlight how to apply for access to the data included in this data note and all other ALSPAC data:

- Please read the ALSPAC access policy, which describes the process of accessing the data and samples in detail, and outlines the costs associated with doing so:
  - http://www.bristol.ac.uk/media-library/sites/alspac/documents/researchers/data-access/ALSPAC_Access_Policy.pdf
- You may also find it useful to browse our fully searchable research proposals database, which lists all research projects that have been approved since April 2011:
  - https://proposals.epi.bristol.ac.uk/?q=proposalSummaries
- Please submit your research proposal for consideration by the ALSPAC Executive Committee.
  - https://proposals.epi.bristol.ac.uk/
- You will receive a response within 10 working days to advise you whether your proposal has been approved.

**MCS:** Millennium Cohort Study data are freely available to bona fide researchers under standard access conditions and can be downloaded on the UK Data Service website: https://ukdataservice.ac.uk/.

**BiB:** The BiB website contains further details about the study, links to detailed data dictionaries, and information about how to apply for access to data: https://borninbradford.nhs.uk/. Researchers are encouraged to make applications to use BiB data. Applications can be made via an expression of interest form available on the study website: https://borninbradford.nhs.uk/research/how-to-access-data/.

