## Supplementary material for "Childhood emotional symptom trajectories in three generationally and socio-ethnically distinct UK birth cohorts"

### Additional methods

#### Details of ethical approval for cohorts

**ALSPAC**: Ethical approval was obtained from the ALSPAC Ethics and Law Committee and the Local Research Ethics Committees. Informed consent for the use of data collected via questionnaires and clinics was obtained from participants following the recommendations of the ALSPAC Ethics and Law Committee at the time. Participants can contact the study team at any time to retrospectively withdraw consent for their data to be used. Study participation is voluntary and during all data collection sweeps, information was provided on the intended use of data.

**MCS**: Data collection was approved by the UK National Health Service Research Ethics Committee. Written consent was obtained from all parents at each survey (for MCS1, South West MREC [MREC/01/6/19]; for MCS2 and MCS3, London MREC [MREC/03/2/022, 05/MRE02/46]; for MCS4, Yorkshire MREC [07/MRE03/32]; for MCS5, Yorkshire and The Humber-Leeds East [11/YH/0203]; for MCS6, London MREC [13/LO/1786]; for MCS7, North East–York [REC ref 17/NE/0341]).

**BiB:** Ethics approval for the Born in Bradford study was granted by the National Health Service Health Research Authority Yorkshire and the Humber (Bradford Leeds) Research Ethics Committee (reference: 07/H1302/112, date of approval 01/04/2008). Informed consent for data collection and linkage to routine healthcare records was provided by participants at recruitment to the BiB cohort study. Further information on our privacy policy can be found here: https://borninbradford.nhs.uk/privacy-policy/.

#### Measurement of maternal anxiety/depression

We used a binary variable for presence/absence of maternal depression and/or anxiety. Maternal depression/anxiety was measured differently across cohorts. In BiB this was measured at baseline using the GHQ-28 screening questionnaire (range 0-84) and dichotomized with scores ≥24 indicating depression/anxiety. In ALSPAC mothers reported any depression or "nerves" (measured when child was 1 year 9 months). In MCS mothers reported at baseline (child 9 months) if they had ever been diagnosed with *serious* depression/anxiety.

#### Additional notes on sample size / power

In latent class analyses, models with more classes, poorer class separation, or missing time points require larger samples. As a rule of thumb, literature suggest a minimum of n=300-500 for 3-4 classes with larger samples of ~1000 cited for 5+ classes (Kim, 2012; Sinha et al., 2020; Tein et al., 2013; Weller et al., 2020; Wurpts & Geiser, 2014).

#### Main models – additional information

We preserved the relative spacing between timepoints and retained a zero anchor. All models were estimated using maximum likelihood which is robust to non-normality. We used 1000 random starts, 200 final iterations. The best log-likelihood was replicated at least 20 times in all models.

#### Missing data

Participants contributed to a model if they had at least one datapoint. Full information maximum likelihood (FIML) was used to fit the model which assumes data are missing at random (MAR) and uses observed information to estimate missing values. Missing data distributions can be found in Tables S2a-2c.

Missing data was <10% for sex and ethnic group in all cohorts (Table S2a). Missing SDQ-E data in ALSPAC and MCS increased with age/time to a maximum level of ~35% at age 12-14yrs in both cohorts. Missingness was <10% for sex and ethnic group in all cohorts (Table S2a). In both MCS and ALSPAC roughly 50% of people had complete data at all four timepoints (Tables S7, S8). In BiB, no participants had data for all timepoints (Tables S6); most commonly people had data at timepoint three (9-11 years; because the Growing Up wave recruited additional participants) or timepoint one (3-5 years; likely due to retention at earlier timepoints) (Table S6).

In all three cohorts there was a trend for mean SDQ score increasing with age category. Mean SDQ scores were similar across cohorts at younger ages (3-8 years). However at 9-11 years, 12-14 years and 15-17 years there were notable differences between cohorts; mean SDQ increased with each generation and scores were highest in the youngest cohort (BiB) at age 12-14.

### Sensitivity analyses

#### Using data up to age 17

We ran supplementary analyses in MCS and ALSPAC using data from age 3-17yrs (Figure S1). When trajectories were extended to include a timepoint at age 15-17yrs in ALSPAC and MCS (Figure S1), the high-level symptom group in ALSPAC continued to increase with symptom scores ~7 points. In MCS the high-level group remained persistent and stable.

**Restricting sample to participants with 2+ repeated measures**

We conducted sensitivity analyses restricting the sample to participants with 2 or more repeated measures to look for any substantial differences in trajectories (Figure S2a). These samples were not considered suitable for primary analyses as the samples were substantially smaller and selection bias is possible (those remaining in the sample at later timepoints may differ systematically from those who lost-to-follow-up). After modelling trajectories we explored missing data patterns to better understand bias in our final chosen models (Tables S6-8).

In sensitivity analyses limiting the sample to 2+ repeated measures, the trajectory groups remained broadly the same in MCS and ALSPAC suggesting minimal bias resulting from missing data. In BiB, the mid-level and low-level trajectories remained broadly the same. However the high-level trajectory changed; the trajectory start point remained high (score of 6) but decreased. This was likely due to data sparsity at the later timepoints and these results are therefore difficult to interpret, hence supplementary (Figure S2a).

**Adjustment of SDQ-E scores based on reporter type in BiB**

We conducted further sensitivity analyses to test whether trajectory groups were an artefact of using SDQ-E scores reported by different reporter types at each timepoint in BiB (Supplementary Material; Table S1b).

First, we calculated mean differences between the observed teacher/self-reports of SDQ-E at each timepoint in the current study, and published parent-reported “norms” at ages corresponding to the timepoints in our study (Table S7). These “norms” are based on a large national survey of child and adolescent mental health carried out by the Office for National Statistics (Meltzer et al., 2000). This representative British sample included 10,438 individuals aged between 5 and 15. Complete SDQ information was obtained from 10,298 parents (99% of sample), 8,208 teachers (79% of sample) and 4,228 11-15 year olds (93% of this age band).

Second, where there were both parent/teacher-or parent/self-reports at a single timepoint in BiB, we calculated mean differences between the teacher/self-reports compared to the parent reported means at the same timepoint and adjusted the non-parent reported scores up or down accordingly (Table S8).

We re-ran three-class LCGA models using each set of adjusted SDQ-E scores, and compared the prevalence, slope and intercepts of the classes to those from the original three-class LCGA model (estimated using observed SDQ-E scores) (Supplementary Figure S1).

**Alternative modelling approaches**

Alternative modelling approaches were considered including Latent Growth Mixture Modelling (LGMM, which allows for within-class variance), and the addition of a quadratic term were considered. Parsimony was a priority given the small sample size in BiB. LCGA was considered a reasonable approach for our objective of identifying broad, interpretable trajectory groups (with a primary interest in a high/persistent class) and comparing these across cohorts.

Figure S2b shows the linear model fits the observed means well therefore suggesting no need for a quadratic model.

#### Table S1a. Average age of participants at each timepoint

| | Mean (SD) age, months<br>Nearest years | | |
|---|---|---|---|
| | **ALSPAC** | **MCS** | **BiB** |
| **3-5yrs<br>(timepoint 1)** | 47.94 (1.47)<br>4 years | 61.07 (7.94)<br>5 years | 61.71 (3.67)<br>5 years |
| **6-8yrs<br>(timepoint 2)** | 95.40 (6.24)<br>8 years | 86.85 (3.07)<br>7 years | 101.50 (4.13)<br>8 years |
| **9-11yrs<br>(timepoint 3)** | 135.94 (9.41)<br>11 years | 128.07 (5.63)<br>11 years | 120.93 (7.82)<br>10 years |
| **12-14yrs<br>(timepoint 4)** | 157.76 (2.32)<br>13 years | 165.01 (5.24)<br>14 years | 167.11 (6.50)<br>14 years |

#### Table S1b. Proportion of reporter type at each timepoint in the Born in Bradford Cohort

| Age | School | Parent | Self | Total | |
|---|---|---|---|---|---|
| **3-5** | 2,167 | | | 2,167 | 100% school/teacher |
| **6-8** | 511 | 861 | | 1,372 | 40% teacher/school, 60% parent |
| **9-11** | 168 | 3,698 | | 3,866 | 4% school, 95% parent |
| **12-14** | | 110 | 1,113 | 1,223 | 9% parent, 91% self-report |

#### Table S2a. Proportion of missing data in baseline sample across all cohorts

| | ALSPAC | MCS | BiB |
|---|---|---|---|
| **Baseline sample** | 11025 (100%) | 15446 (100%) | 6711 (100%) |
| **Sex (at birth)** | | | |
| % Female (at birth) | 5631 (51%) | 7566 (49%) | 3278 (49%) |
| % Male (at birth) | 5373 (49%) | 7252 (47%) | 3433 (51%) |
| Missing | 21 (0.19%) | 628 (4%) | 0 (0%) |
| **Ethnic group** | | | |
| **White:** English, Welsh, Scottish, Northern Irish or British; Irish; any other White background | 9533 (86%) | 12,287 (80%) | 2288 (34%) |
| **Ethnic minority groups:** Mixed/multiple, Asian or Asian British, Black, Black British, Caribbean or African, Arab, Other ethnic group | 410 (4%) | 2496 (16%) | 4422 (66%) |
| Missing | 1082 (10%) | 663 (4%) | 1 (0.01%) |
| **Mother's highest education** | | | |
| <5 GCSEs/Other/Unknown | 2676 (24%) | 4727 (31%) | 1677 (25%) |
| GCSEs | 3603 (33%) | 4925 (32%) | 1715 (26%) |
| A level | 2437 (22%) | 1413 (9%) | 826 (12%) |
| Degree/higher | 1436 (13%) | 3706 (24%) | 1391 (21%) |
| Missing | 873 (8%) | 675 (4%) | 1102 (16%) |
| **Home ownership status** | | | |
| Owns/mortgage | 8029 (73%) | 8383 (54%) | 4059 (60%) |
| Private rent | 637 (6%) | 1198 (8%) | 888 (13%) |
| Social/council | 1280 (12%) | 3784 (25%) | 537 (8%) |
| Other/unknown | 304 (3%) | 905 (6%) | 128 (2%) |
| Missing | 775 (7%) | 1176 (8%) | 1099 (16%) |
| **IMD 2010 quintile** | | | |
| Q1 | 854 (8%) | 3194 (21%) | 3822 (57%) |
| Q2 | 1138 (10%) | 1978 (13%) | 985 (15%) |
| Q3 | 1117 (10%) | 1567 (11%) | 605 (9%) |
| Q4 | 1446 (13%) | 1260 (8%) | 137 (2%) |
| Q5 | 1674 (15%) | 1147 (7%) | 69 (1%) |
| Missing | 4796 (44%) | 6300 (41%) | 1093 (16%) |
| **Maternal dep/anx** | | | |
| No | 8180 (74%) | 11,168 (72%) | 2683 (40%) |
| Yes | 2845 (26%) | 3623 (23%) | 2054 (31%) |
| Missing | 0 (0%) | 655 (4%) | 1974 (29%) |
| **SDQ-E score** | | | |
| 3-5 years | 9166 (83%) | 14,619 (95%) | 2167 (32%) |
| Missing | 1859 (17%) | 827 (5%) | 4544 (68%) |
| 6-8 years | 8995 (82%) | 12,055 (78%) | 1372 (20%) |
| Missing | 2030 (18%) | 3391 (22%) | 5339 (80%) |
| 9-11 years | 8606 (78%) | 11417 (74%) | 3866 (58%) |
| Missing | 2419 (22%) | 4029 (26%) | 2845 (42%) |
| 12-14 years | 7039 (64%) | 10,021 (65%) | 1223 (18%) |
| Missing | 3986 (36%) | 5425 (35%) | 5488 (82%) |
| 15-17 years | 3618 (33%) | 2412 (16%) | 4 (0.01%) |
| Missing | 7407 (67%) | 13034 (84%) | 6707 (99.9%) |

**Table S2b. Predictors of missingness from analysis sample**

| | | | Included in the derivation of trajectories? (1 or more repeated measure) | | | | | | | | |
|---|---|---|---|---|---|---|---|---|---|---|---|
| | | | *ALSPAC* | | | *MCS* | | | *BiB* | | |
| **Measure** | | | **Yes** | **No** | ***P*** | **Yes** | **No** | ***P*** | **Yes** | **No** | ***P*** |
| **Sex** | **male** | **N (%)** | 5631 (51) | 1944 (51) | 0.66 | 7566 (51) | 1129 (54) | 0.009 | 3433 (51) | 3720 (52) | 0.288 |
| | **female** | **N (%)** | 5373 (49) | 1886 (49) | | 7252 (49) | 957 (46) | | 3278 (49) | 3426 (48) | |
| **Ethnic group** | **White** | **N (%)** | 9533 (96) | 1836 (90) | <0.0001 | 12,287 (83) | 1589 (77) | <0.0001 | 2288 (34) | 3155 (44) | <0.0001 |
| | **Not white** | **N (%)** | 410 (4) | 193 (10) | | 2496 (17) | 485 (23) | | 4422 (66) | 3956 (56) | |
| **IMD (quintiles)** | **1 (most deprived)** | **N (%)** | 854 (14) | 478 (26) | <0.0001 | 3194 (35) | 893 (38) | 0.05 | 3822 (68) | 3032 (64) | <0.0001 |
| | **2** | **N (%)** | 1138 (18) | 415 (22) | | 1978 (22) | 518 (22) | | 985 (18) | 899 (19) | |
| | **3** | **N (%)** | 1117 (18) | 306 (16) | | 1567 (17) | 409 (17) | | 605 (11) | 559 (12) | |
| | **4** | **N (%)** | 1446 (23) | 350 (19) | | 1260 (14) | 300 (13) | | 137 (2) | 161 (3) | |
| | **5 (least deprived)** | **N (%)** | 1674 (27) | 317 (17) | | 1147 (13) | 260 (11) | | 69 (1) | 104 (2) | |
| **Maternal depression/anxiety** | **No** | **N (%)** | 8180 (74) | 4275 (93) | <0.0001 | 11,168 (76) | 1605 (77) | 0.07 | 2683 (57) | 2130 (56) | 0.37 |
| | **Yes** | **N (%)** | 2845 (26) | 345 (7) | | 3623 (24) | 471 (23) | | 2054 (43) | 1696 (44) | |

### Table S2c. Distribution of missing data in ALSPAC age 24 and age 30 depression/GAD diagnosis variables in analysis sample

| | | | Included (not missing) in association between latent classes and depression diagnosis (any severity) | | | | | | Included (not missing) in association between latent classes and GAD diagnosis | | | | | |
|---|---|---|---|---|---|---|---|---|---|---|---|---|---|---|
| | | | Age 24 | | | Age 30 | | | Age 24 | | | Age 30 | | |
| **Measure** | | | **No** | **Yes** | ***P*** | **No** | **Yes** | ***P*** | **No** | **Yes** | ***P*** | **No** | **Yes** | ***P*** |
| **SDQ-E trajectory group** | **High-symptom** | **N (%)** | 418 (6) | 161 (4) | <0.001 | 413 (5) | 166 (5) | 0.157 | 419 (6) | 160 (4) | 0.001 | 1457 (19) | 684 (20) | 0.16 |
| | **Mid-symptom** | **N (%)** | 1384 (19) | 757 (21) | | 1454 (19) | 687 (20) | | 1386 (19) | 755 (21) | | 5800 (76) | 2505 (75) | |
| | **Low-symptom** | **N (%)** | 5606 (76) | 2699 (75) | | 5793 (76) | 2512 (75) | | 5612 (76) | 2693 (75) | | 414 (5) | 165 (5) | |
| **Sex** | **male** | **N (%)** | 4259 (58) | 1372 (38) | <0.001 | 4320 (57) | 1311 (39) | <0.001 | 4262 (58) | 1369 (38) | <0.001 | 4323 (57) | 1308 (39) | <0.001 |
| | **female** | **N (%)** | 3128 (42) | 2245 (62) | | 3319 (43) | 2054 (61) | | 3134 (42) | 2239 (62) | | 3327 (43) | 2046 (61) | |
| **IMD (quintiles)** | **1 (most deprived)** | **N (%)** | 676 (16) | 178 (9) | <0.001 | 688 (16) | 166 (9) | <0.001 | 676 (16) | 178 (9) | 0.001 | 689 (16) | 165 (9) | <0.001 |
| | **2** | **N (%)** | 842 (20) | 296 (15) | | 833 (19) | 305 (17) | | 843 (20) | 295 (15) | | 834 (19) | 304 (17) | |
| | **3** | **N (%)** | 771 (18) | 346 (17) | | 799 (18) | 318 (17) | | 775 (18) | 342 (17) | | 801 (18) | 316 (17) | |
| | **4** | **N (%)** | 917 (22) | 529 (27) | | 966 (22) | 480 (26) | | 917 (22) | 529 (27) | | 966 (22) | 48 (26) | |
| | **5 (least deprived)** | **N (%)** | 1044 (25) | 630 (32) | | 1104 (25) | 570 (31) | | 1046 (25) | 628 (32) | | 1106 (25) | 568 (31) | |
| **Maternal depression/anxiety** | **No** | **N (%)** | 5438 (73) | 2742 (76) | 0.007 | 5648 (74) | 2532 (75) | 0.10 | 5445 (73) | 2735 (76) | 0.007 | 5653 (74) | 2527 (75) | 0.07 |
| | **Yes** | **N (%)** | 1970 (27) | 875 (24) | | 2012 (26) | 833 (25) | | 1972 (27) | 873 (24) | | 2018 (26) | 827 (25) | |

**Table S3. Fit statistics for 1-5 class models: BiB cohort**

| Number of classes | Class N, (%) (estimated posterior probabilities) | Class average posterior probability | AIC | BIC | SABIC | Entropy | Bootstrap LRT 2 * log-likelihood diff, p -value | LMR-LRT; p-value |
|---|---|---|---|---|---|---|---|---|
| 1 | 1 - 6711 (100%) | | 37244 | 37284.87 | 37265.8 | n/a | | |
| 2 | 1 - 5788 (86%)<br>2 - 922 (14%) | 1 - 95%<br>2 - 81% | 36084.63 | 36145.93 | 36117.33 | 0.75 | 1165.37; p=0.0000 | 1122.891; p=0.001 |
| 3 | 1 - 498 (7%)<br>2 - 1179 (18%)<br>3 - 5032 (75%) | 1 - 80%<br>2 - 89%<br>3 - 85% | 35420.68 | 35502.42 | 35464.29 | 0.64 | 669.947; p=0.0000 | 645.527; p=0.005 |
| 4 | 1 - 1023 (15%)<br>2 - 305 (5%)<br>3 - 4907 (73%)<br>4 - 476 (7%) | 1 - 98%<br>2 - 77%<br>3 - 83%<br>4 - 80% | 34914.28 | 35016.45 | 34968v79 | 0.66 | 512.405; p=0.0000 | 493.728; p=0.48 |
| 5 | 1 - 486 (7%)<br>2 - 4439 (66%)<br>3 - 205 (3%)<br>4 - 998 (15%)<br>5 - 583 (9%) | 1 - 80%<br>2 - 79%<br>3 - 79%<br>4 - 84%<br>5 - 69% | 34716.06 | 34838.67 | 34781.47 | 0.63 | 204.218; p=0.0000 | 196.774; p=0v002 |
| 6 | 1 - 446 (7%)<br>2 - 746 (11%)<br>3 - 986 (15%)<br>4 - 148 (2%)<br>5 - 4264 (64%)<br>6 - 122 (2%) | 1 - 75%<br>2 - 58%<br>3 - 95%<br>4 - 78%<br>5 - 79%<br>6 - 98% | 34499.36 | 34642.4 | 34575.67 | 0.64 | 187.358; p=0.0000 | 180.529; p=0.008 |

**Table S4. Fit statistics, 1-5 class models: MCS cohort**

| Number of classes | Class N, (%) (estimated posterior probabilities) | Class average posterior probability | AIC | BIC | SABIC | Entropy | BLRT | LMR-LRT; p-value |
|---|---|---|---|---|---|---|---|---|
| 1 | 100% | 100% | 195454.723 | 195500.6 | 195481.5 | n/a | n/a | n/a |
| 2 | 1 - 12735 (82%)<br>2 - 2711 (18%) | 1 - 97%<br>2 - 90% | 183847.545 | 183916.4 | 183887.8 | 0.85 | -97721.362; p=0.0000 | 1225.235; p=0.0000 |
| 3 | 1 - 3315 (21%)<br>2 - 896 (6%)<br>3 - 11234 (73%) | 1 - 80%<br>2 - 88%<br>3 - 94% | 181592.083 | 181683.8 | 181645.7 | 0.79 | -91914.773; p=0.0000 | 2185.917; p=0.0000 |
| 4 | 1 - 10880 (70%)<br>2 - 664 (4%)<br>3 - 1865 (12%)<br>4 - 2036 (13%) | 1 - 93%<br>2 - 89%<br>3 - 81%<br>4 - 79% | 179476.398 | 179591.1 | 179543.4 | 0.81 | -90784.042; p=0.0000 | 2050.81; p=0.0013 |
| 5 | 1 - 727 (5%)<br>2 - 9941 (64%)<br>3 - 1989 (13%)<br>4 - 2278 (15%)<br>5 - 511 (3%) | 1 - 81%<br>2 - 90%<br>3 - 78%<br>4 - 75%<br>5 - 89% | 178510.542 | 178648.2 | 178591 | 0.78 | -89723.199; p=0.0000 | 939.390; p=0.006 |
| 6 | 1 - 2023 (13%)<br>2 - 9538 (62%)<br>3 - 693 (4%)<br>4 - 161 (1%)<br>5 - 752 (5%)<br>6 - 2280 (15%) | 1 - 76%<br>2 - 89%<br>3 - 80%<br>4 - 86%<br>5 - 82%<br>6 - 75% | 177906.594 | 178067.1 | 178000.4 | 0.78 | -89237.271; p=0.0000 | 589.573; p=0.06 |

**Table S5. Fit statistics, 1-5 class models: ALSPAC cohort**

| Number of classes | Class N (estimated posterior probabilities) | Class average posterior probability | AIC | BIC | SABIC | Entropy | BLRT | LMR-LRT; p-value |
|---|---|---|---|---|---|---|---|---|
| 1 | 100% | 100% | 131720.4 | 131764.2 | 131745.2 | n/a | | |
| 2 | 1 - 2027 (18%)<br>2 - 8998 (82%) | 1 - 89%<br>2 - 96% | 124156.5 | 124222.3 | 124193.7 | 0.82 | -65854.186; p=0.0000 | 7308.157; p=0.0000 |
| 3 | 1 - 2382 (22%)<br>2 - 7990 (72%)<br>3 - 653 (6%) | 1 - 81%<br>2 - 93%<br>3 - 86% | 122494.7 | 122582.4 | 122544.3 | 0.79 | -62069.248; p=0.0000 | 1610.13; p=0.0000 |
| 4 | 1 - 1040 (9%)<br>2 - 1766 (16%)<br>3 - 7607 (69%)<br>4 - 611 (6%) | 1 - 76%<br>2 - 79%<br>3 - 91%<br>4 - 84% | 121276.8 | 121386.4 | 121338.7 | 0.78 | -61235.352; p=0.0000 | 1181.615; p=0.0000 |
| 5 | 1 - 1759 (16%)<br>2 - 7082 (64%)<br>3 - 405 (4%)<br>4 - 405 (4%)<br>5 - 1374 (12%) | 1 - 79%<br>2 - 90%<br>3 - 81%<br>4 - 83%<br>5 - 73% | 120419.7 | 120551.2 | 120494 | 0.78 | -60623.387; p=0.0000 | 833.256; p=0.0000 |
| 6 | 1 - 275 (2%)<br>2 - 1964 (18%)<br>3 - 720 (7%)<br>4 - 1187 (11%)<br>5 - 400 (4%)<br>6 - 6479 (59%) | 1 - 83%<br>2 - 74%<br>3 - 63%<br>4 - 69%<br>5 - 72%<br>6 - 95% | 119893.3 | 120046.7 | 119980 | 0.75 | -60191.839; p=0.0000 | 514.017; p=0.0001 |

**Table S6. Results of pairwise Wald tests comparing slopes and intercepts of trajectory classes across cohorts**

| Trajectory class | Pairwise comparison | Parameter | Wald χ² | df | p-value | raw P | Adjusted for multiple testing |
|---|---|---|---|---|---|---|---|
| High | ALSPAC vs MCS | Slope | 27.635 | 1 | p<0.001 | 1.4650E-07 | 4.39511E-07 |
| High | ALSPAC vs BIB | Slope | 33.587 | 1 | p<0.001 | 6.81467E-09 | 2.0444E-08 |
| High | MCS vs BIB | Slope | 0.004 | 1 | 0.9526 | 0.949570971 | 2.848712913 |
| High | ALSPAC vs MCS | Intercept | 75.909 | 1 | p<0.001 | 2.97043E-18 | 8.91129E-18 |
| High | ALSPAC vs BIB | Intercept | 101.563 | 1 | p<0.001 | 6.92264E-24 | 2.07679E-23 |
| High | MCS vs BIB | Intercept | 8.605 | 1 | 0.0034 | 0.003352414 | 0.010057241 |
| Mid | ALSPAC vs MCS | Slope | 40.748 | 1 | p<0.001 | 1.73181E-10 | 5.19542E-10 |
| Mid | ALSPAC vs BIB | Slope | 0.288 | 1 | 0.5915 | 0.591505037 | 1.774515111 |
| Mid | MCS vs BIB | Slope | 7.74 | 1 | 0.0054 | 0.00540108 | 0.01620324 |
| Mid | ALSPAC vs MCS | Intercept | 250.573 | 1 | p<0.001 | 1.9477E-56 | 5.84309E-56 |
| Mid | ALSPAC vs BIB | Intercept | 1134.941 | 1 | p<0.001 | 8.4094E-249 | 2.5228E-248 |
| Mid | MCS vs BIB | Intercept | 434.587 | 1 | p<0.001 | 1.63132E-96 | 4.89397E-96 |
| Low | ALSPAC vs MCS | Slope | 53.492 | 1 | p<0.001 | 2.59648E-13 | 7.78944E-13 |
| Low | ALSPAC vs BIB | Slope | 137.468 | 1 | p<0.001 | 9.52661E-32 | 2.85798E-31 |
| Low | MCS vs BIB | Slope | 53.988 | 1 | p<0.001 | 2.01718E-13 | 6.05153E-13 |
| Low | ALSPAC vs MCS | Intercept | 273.602 | 1 | p<0.001 | 1.86161E-61 | 5.58482E-61 |
| Low | ALSPAC vs BIB | Intercept | 226.357 | 1 | p<0.001 | 3.71405E-51 | 1.11421E-50 |
| Low | MCS vs BIB | Intercept | 22.795 | 1 | p<0.001 | 1.80234E-06 | 5.40702E-06 |

**Table S7. Parent Teacher and Self-reported mean (SD) of SDQ – national normative values for emotional subscale**

| | Parent | Teacher | Self | Difference |
|---|---|---|---|---|
| **5-10 years** | 1.9 (2.0) | 1.5 (1.9) | Not available, too young | Parents over-estimate by 0.40 vs teachers |
| **11-15 years** | 1.9 (2.0) | 1.3 (1.9) | 2.8 (2.1) | Parents over-estimate by 0.60 vs teachers<br>YP over-estimate by 1.5 vs teachers<br>YP over-estimate by 0.9 vs parents |

**Adjustments made based on this table:**

*at 3-5 adjust teacher by + 0.40

*at 6-8 adjust teacher by +0.40

*at 9-11 +0.5 to teacher reports – this timepoint bridges the 5-10 and 11-15 age bands in ONS study, so I took 0.50 as mid-point

*at 12-14 -0.9 from YP reports

**Table S8. Mean SDQ-E score differences between reporter types observed within the BiB cohort**

| | SDQ-E score by reporter type: observed in BiB cohort | | | | |
|---|---|---|---|---|---|
| | **Teacher** | **Parent** | **Self** | **Mean difference** | **Description** |
| **3-5** | 1.05 (1.65); n=2167 | - | - | | |
| **6-8** | 1.51 (1.91), n=511 | 2.23 (2.16), n=861 | - | 0.72 | Parent over reports by 0.72 vs teacher |
| **9-11** | 1.63 (2.10), n=168 | 2.24 (2.17), n=3698 | - | 0.61 | Parent over reports by 0.61 vs teacher |
| **12-14** | - | 2.01 (2.43), n=110 | 3.87 (2.71), n=1113 | 1.86 | YPs over report by 1.86 |

**Adjustments were as follows:**

*age 3-5, added 0.67 to all reports: based on midpoint between difference at age 6-8 (0.72) and 9-11 (0.61) as all teacher reports at age 3-5

*age 6-8, added 0.72 to teacher reports

*9-11, added 0.61 to teacher reports

*12-14, subtracted 1.86 from self-reports

**Table S9. BiB overall missing data pattern: 3 class trajectory model**

| Pattern | Frequency | Percent (%) |
|---|---|---|
| * * 1 * | 2,464 | 36.72 |
| 1 * * * | 1,035 | 15.42 |
| 1 * 1 * | 972 | 14.48 |
| * 1 * * | 754 | 11.24 |
| * * * 1 | 646 | 9.63 |
| * 1 * 1 | 317 | 4.72 |
| * * 1 1 | 207 | 3.08 |
| * 1 1 * | 118 | 1.76 |
| 1 1 * * | 79 | 1.18 |
| 1 1 1 * | 66 | 0.98 |
| * 1 1 1 | 38 | 0.57 |
| 1 * * 1 | 14 | 0.21 |
| 1 * 1 1 | 1 | 0.01 |
| Total | 6,711 | 100 |

**Table S10. MCS overall missing data pattern: 3 class trajectory model**

| Pattern | Frequency | Percent (%) |
|---|---|---|
| 1 1 1 1 | 8,111 | 52.51 |
| 1 * * * | 1,819 | 11.78 |
| 1 1 1 * | 1,705 | 11.04 |
| 1 1 * * | 1,225 | 7.93 |
| 1 * 1 1 | 626 | 4.05 |
| 1 1 * 1 | 556 | 3.6 |
| 1 * 1 * | 392 | 2.54 |
| * 1 1 1 | 213 | 1.38 |
| * * 1 1 | 194 | 1.26 |
| 1 * * 1 | 185 | 1.2 |
| * 1 * * | 108 | 0.7 |
| * * 1 * | 94 | 0.61 |
| * 1 1 * | 82 | 0.53 |
| * * * 1 | 81 | 0.52 |
| * 1 * 1 | 55 | 0.36 |
| Total | 15,446 | 100 |

**Table S11. ALSPAC overall missing data pattern: 3 class trajectory model**

| Pattern | Frequency | Percent (%) |
|---|---:|---:|
| 1 1 1 1 | 5,543 | 50.28 |
| 1 1 1 * | 1,259 | 11.42 |
| 1 * * * | 879 | 7.97 |
| 1 1 * * | 805 | 7.3 |
| * 1 1 1 | 543 | 4.93 |
| * * 1 1 | 336 | 3.05 |
| * 1 * * | 307 | 2.78 |
| * 1 1 * | 272 | 2.47 |
| * * 1 * | 267 | 2.42 |
| 1 1 * 1 | 217 | 1.97 |
| 1 * 1 * | 197 | 1.79 |
| 1 * 1 1 | 189 | 1.71 |
| * * * 1 | 85 | 0.77 |
| 1 * * 1 | 77 | 0.7 |
| * 1 * 1 | 49 | 0.44 |
| Total | 11,025 | 100 |

**Table S12. Associations between sociodemographic factors and trajectory groups in ALSPAC**

| Odds of belonging to high and mid symptom trajectories (vs low) by different sociodemographic factors | | | | |
|---|---|---|---|---|
| **ALSPAC (n=11,025)** | **Analysis sample size (CCA)** | **Low (72%) REFERENCE** | **Mid (22%) OR vs ref** | **High (6%) OR vs ref** |
| **Female (at birth)** | 11004 | 1.00 (ref) | **1.59 (1.39-1.80)** | **1.44 (1.18-1.75)** |
| **Ethnic group** | 9943 | 1.00 (ref) | 1.07 (0.77-1.49) | 1.15 (0.69-1.89) |
| **IMD quintile** | | c2 ref | c1 | c3 |
| Q1 - most deprived | | 1.00 (ref) | 1.00 (ref) | 1.00 (ref) |
| Q2 | 1992 | 1.00 (ref) | 0.84 (0.62-1.13) | 0.59 (0.37-0.92) |
| Q3 | 1971 | 1.00 (ref) | 0.92 (0.68-1.24) | 0.69 (0.45-1.07) |
| Q4 | 2300 | 1.00 (ref) | 0.76 (0.57-1.01) | **0.65 (0.43-0.97)** |
| Q5 - least deprived | 2528 | 1.00 (ref) | 0.82 (0.62-1.08) | **0.52 (0.34-0.79)** |
| **Maternal depression/anxiety** | | | | |
| Depression or anxiety | 11,025 | 1.00 (ref) | **1.61 (1.40-1.85)** | **2.54 (2.08-3.10)** |

**Table S13. Associations between sociodemographic factors and trajectory groups in MCS**

| Odds of belonging to high and mid symptom trajectories (vs low) by different sociodemographic factors | | | | |
|---|---|---|---|---|
| **MCS (n=15,446)** | **Analysis sample size (CCA)** | **LOW (73%) REFERENCE** | **MID (21%) OR vs ref** | **HIGH (6%) OR vs ref C2** |
| **Female (at birth)** | 14,818 | 1.00 (ref) | **1.25 (1.12-1.40)** | 1.09 (0.92-1.28) |
| **Ethnicity** | | | | |
| White | | 1.00 (ref) | 1.00 (ref) | 1.00 (ref) |
| Non-white | 14783 | 1.00 (ref) | **1.27 (1.10-1.47)** | **1.92 (1.59-2.32)** |
| **IMD quintile** | | c3 | c1 | c2 |
| Q1 - most deprived | | 1.00 (ref) | 1.00 (ref) | 1.00 (ref) |
| Q2 | 5172 | 1.00 (ref) | 0.86 (0.71-1.03) | **0.62 (0.48-0.80)** |
| Q3 | 4761 | 1.00 (ref) | **0.71 (0.58 -0.87)** | **0.45 (0.33-0.62)** |
| Q4 | 4454 | 1.00 (ref) | **0.54 (0.43-0.69)** | **0.32 (0.22-0.47)** |
| Q5 - least deprived | 4341 | 1.00 (ref) | **0.51 (0.40-0.65)** | **0.26 (0.17-0.41)** |
| **Maternal depression/anxiety** | | | | |
| Has depression/anxiety | 14791 | 1.00 (ref) | **1.74 (1.54-1.96)** | **2.57 (2.17-3.04)** |

**Table S14. Associations between sociodemographic factors and trajectory groups in BiB**

| Odds of belonging to high and mid symptom trajectories (vs low) by different sociodemographic factors | | | | |
|---|---|---|---|---|
| BiB (n=6711) | **Analysis sample size (CCA)** | **LOW (75%) REFERENCE** | **MID (18%)** | **HIGH (7%) OR vs ref** |
| **Female (at birth)** | 6711 | 1.00 (ref) | **1.30 (1.01-1.67)** | **1.45 (1.12-1.89)** |
| **Ethnic group** | | | | |
| White | 6710 | 1.00 (ref) | 1.00 (ref) | 1.00 (ref) |
| Non-white | 6710 | 1.00 (ref) | **0.76 (0.59-0.99)** | **0.70 (0.53-0.91)** |
| **IMD quintile** | | | | |
| Q1 (most deprived) | | 1.00 (ref) | 1.00 (ref) | 1.00 (ref) |
| Q2 | 4807 | 1.00 (ref) | 1.22 (0.84-1.79) | 0.92 (0.54-1.55) |
| Q3 | 4427 | 1.00 (ref) | 0.97 (0.62-1.51) | 0.87 (0.54-1.41) |
| Q4 | 3959 | 1.00 (ref) | 0.38 (0.11-1.37) | 0.67 (0.25-1.80) |
| Q5 (least deprived) | 3891 | 1.00 (ref) | 0.52 (0.11-2.61) | 1.37 (0.50-3.81) |
| **Maternal depression/anxiety** | | | | |
| GHQ-28 score 24 or more | 4737 | 1.00 (ref) | 1.20 (0.88-1.62) | **1.67 (1.23-2.27)** |

**Table S15. Numerical description of 3-class solution and 95% confidence interval calculations**

| cohort | trajectory | timepoint | SDQ-E score | lower | upper | se |
|---|---|---|---|---|---|---|
| BiB (Born: 2007-10) | High | 0 | 5.97 | 5.59368 | 6.34632 | 0.192 |
| BiB (Born: 2007-10) | High | 1 | 6.01 | 5.65132 | 6.36868 | 0.183 |
| BiB (Born: 2007-10) | High | 2 | 6.05 | 5.57176 | 6.52824 | 0.244 |
| BiB (Born: 2007-10) | High | 3 | 6.09 | 5.42556 | 6.75444 | 0.339 |
| BiB (Born: 2007-10) | Mid | 0 | 2.87 | 2.65636 | 3.08364 | 0.109 |
| BiB (Born: 2007-10) | Mid | 1 | 2.67 | 2.474 | 2.866 | 0.1 |
| BiB (Born: 2007-10) | Mid | 2 | 2.47 | 2.18188 | 2.75812 | 0.147 |
| BiB (Born: 2007-10) | Mid | 3 | 2.27 | 1.84664 | 2.69336 | 0.216 |
| BiB (Born: 2007-10) | Low | 0 | 0.27 | 0.21512 | 0.32488 | 0.028 |
| BiB (Born: 2007-10) | Low | 1 | 1.15 | 1.09904 | 1.20096 | 0.026 |
| BiB (Born: 2007-10) | Low | 2 | 2.03 | 1.9418 | 2.1182 | 0.045 |
| BiB (Born: 2007-10) | Low | 3 | 2.91 | 2.77476 | 3.04524 | 0.069 |
| ALSPAC (Born: 1990-92) | Mid | 0 | 2.51 | 2.3532 | 2.6668 | 0.08 |
| ALSPAC (Born: 1990-92) | Mid | 1 | 2.687 | 2.5596 | 2.8144 | 0.065 |
| ALSPAC (Born: 1990-92) | Mid | 2 | 2.864 | 2.72288 | 3.00512 | 0.072 |
| ALSPAC (Born: 1990-92) | Mid | 3 | 3.041 | 2.85284 | 3.22916 | 0.096 |
| ALSPAC (Born: 1990-92) | Low | 0 | 1.099 | 1.05588 | 1.14212 | 0.022 |
| ALSPAC (Born: 1990-92) | Low | 1 | 0.957 | 0.92368 | 0.99032 | 0.017 |
| ALSPAC (Born: 1990-92) | Low | 2 | 0.815 | 0.78364 | 0.84636 | 0.016 |
| ALSPAC (Born: 1990-92) | Low | 3 | 0.673 | 0.6338 | 0.7122 | 0.02 |
| ALSPAC (Born: 1990-92) | High | 0 | 3.469 | 3.15932 | 3.77868 | 0.158 |
| ALSPAC (Born: 1990-92) | High | 1 | 4.396 | 4.16864 | 4.62336 | 0.116 |
| ALSPAC (Born: 1990-92) | High | 2 | 5.323 | 5.05252 | 5.59348 | 0.138 |
| ALSPAC (Born: 1990-92) | High | 3 | 6.25 | 5.85212 | 6.64788 | 0.203 |
| MCS (Born: 2000-02) | Mid | 0 | 2.324 | 2.15936 | 2.48864 | 0.084 |
| MCS (Born: 2000-02) | Mid | 1 | 2.905 | 2.80112 | 3.00888 | 0.053 |
| MCS (Born: 2000-02) | Mid | 2 | 3.486 | 3.32136 | 3.65064 | 0.084 |
| MCS (Born: 2000-02) | Mid | 3 | 4.067 | 3.79064 | 4.34336 | 0.141 |
| MCS (Born: 2000-02) | High | 0 | 5.283 | 5.01644 | 5.54956 | 0.136 |
| MCS (Born: 2000-02) | High | 1 | 5.338 | 5.13808 | 5.53792 | 0.102 |
| MCS (Born: 2000-02) | High | 2 | 5.393 | 4.99904 | 5.78696 | 0.201 |
| MCS (Born: 2000-02) | High | 3 | 5.448 | 4.80316 | 6.09284 | 0.329 |
| MCS (Born: 2000-02) | Low | 0 | 0.813 | 0.77184 | 0.85416 | 0.021 |
| MCS (Born: 2000-02) | Low | 1 | 0.942 | 0.91064 | 0.97336 | 0.016 |
| MCS (Born: 2000-02) | Low | 2 | 1.071 | 1.03572 | 1.10628 | 0.018 |
| MCS (Born: 2000-02) | Low | 3 | 1.2 | 1.151 | 1.249 | 0.025 |

**Figure S1. Comparison of original BiB cohort three class model vs. models fit with SDQ-E mean scores adjusted to account for differences in reporter type across timepoints**

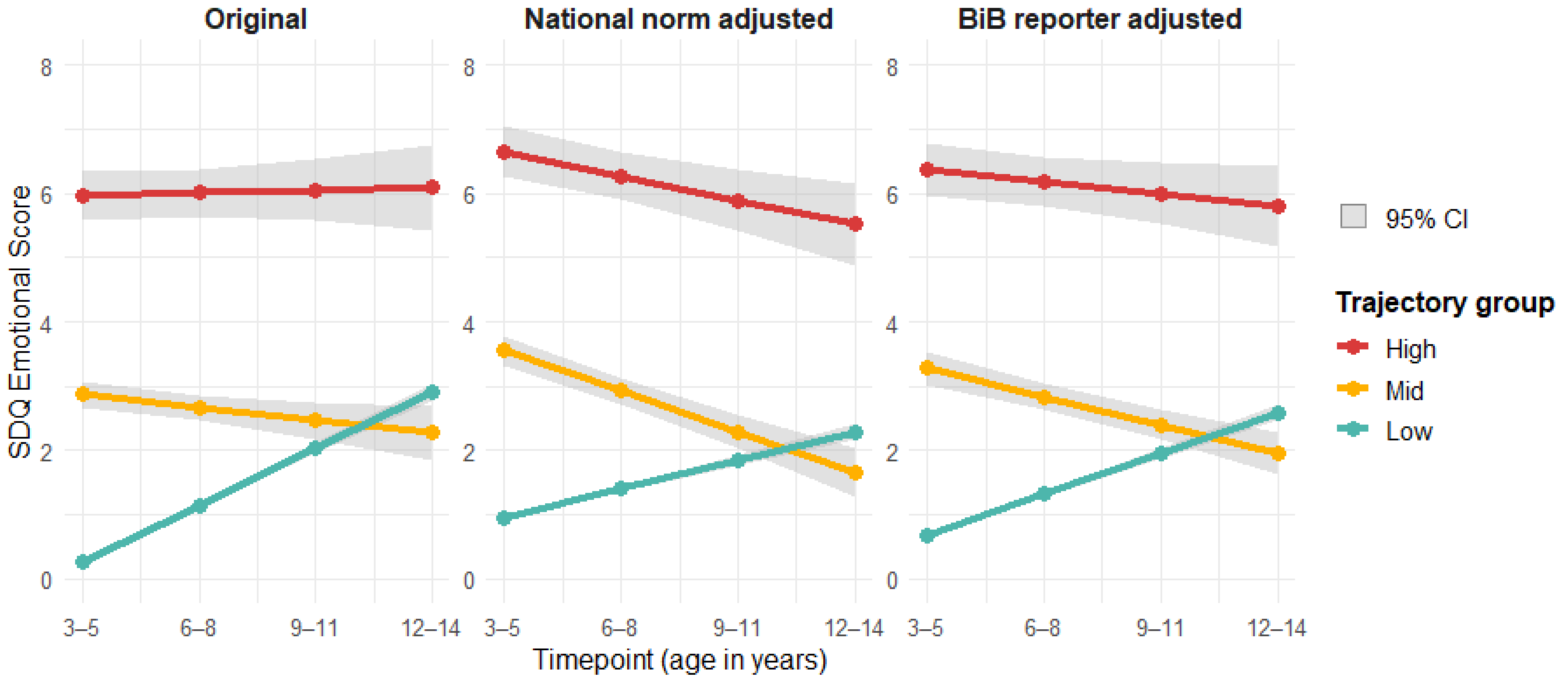

Footnote: confidence intervals are present for the low symptom trajectory group. However, these are very small due to the large sample in this group.

**Figure S2. Trajectories of emotional symptoms from age 3-17 years in ALSPAC and MCS**

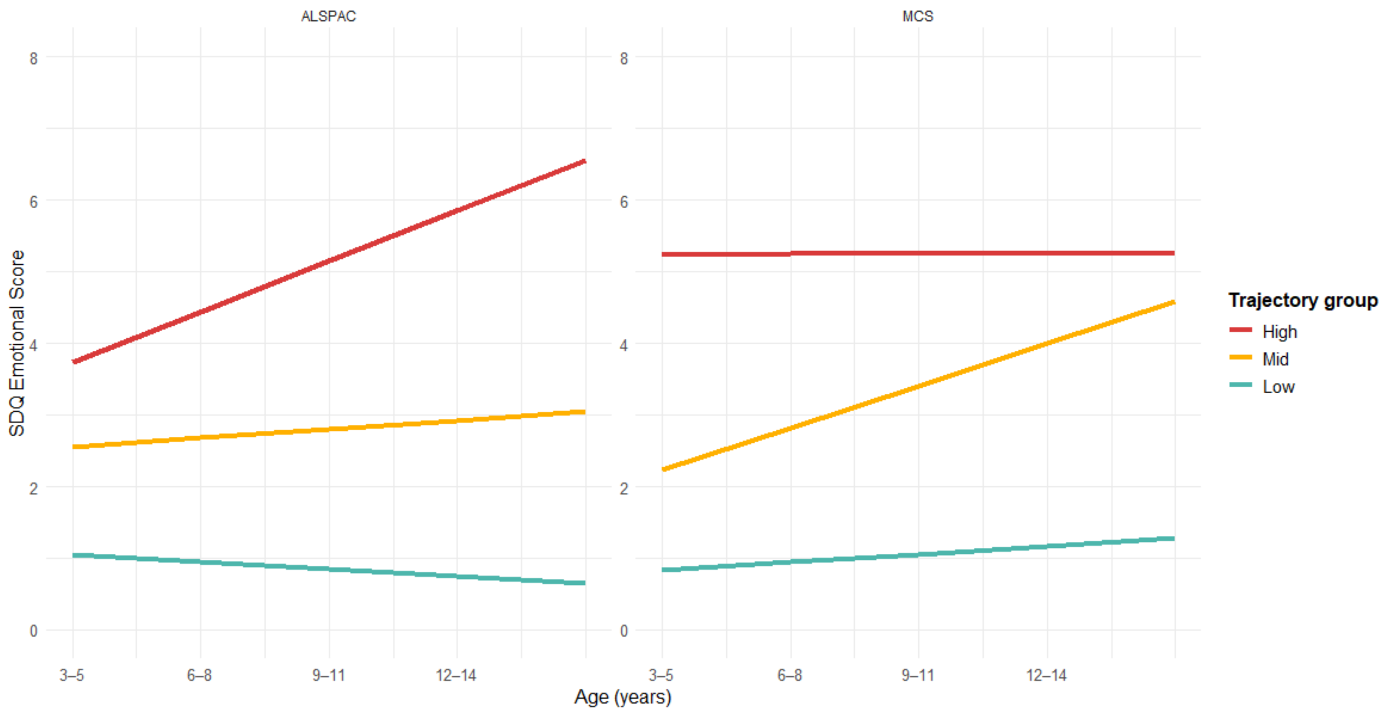

**Figure S3a. Sample restricted to 2+ repeated measures**

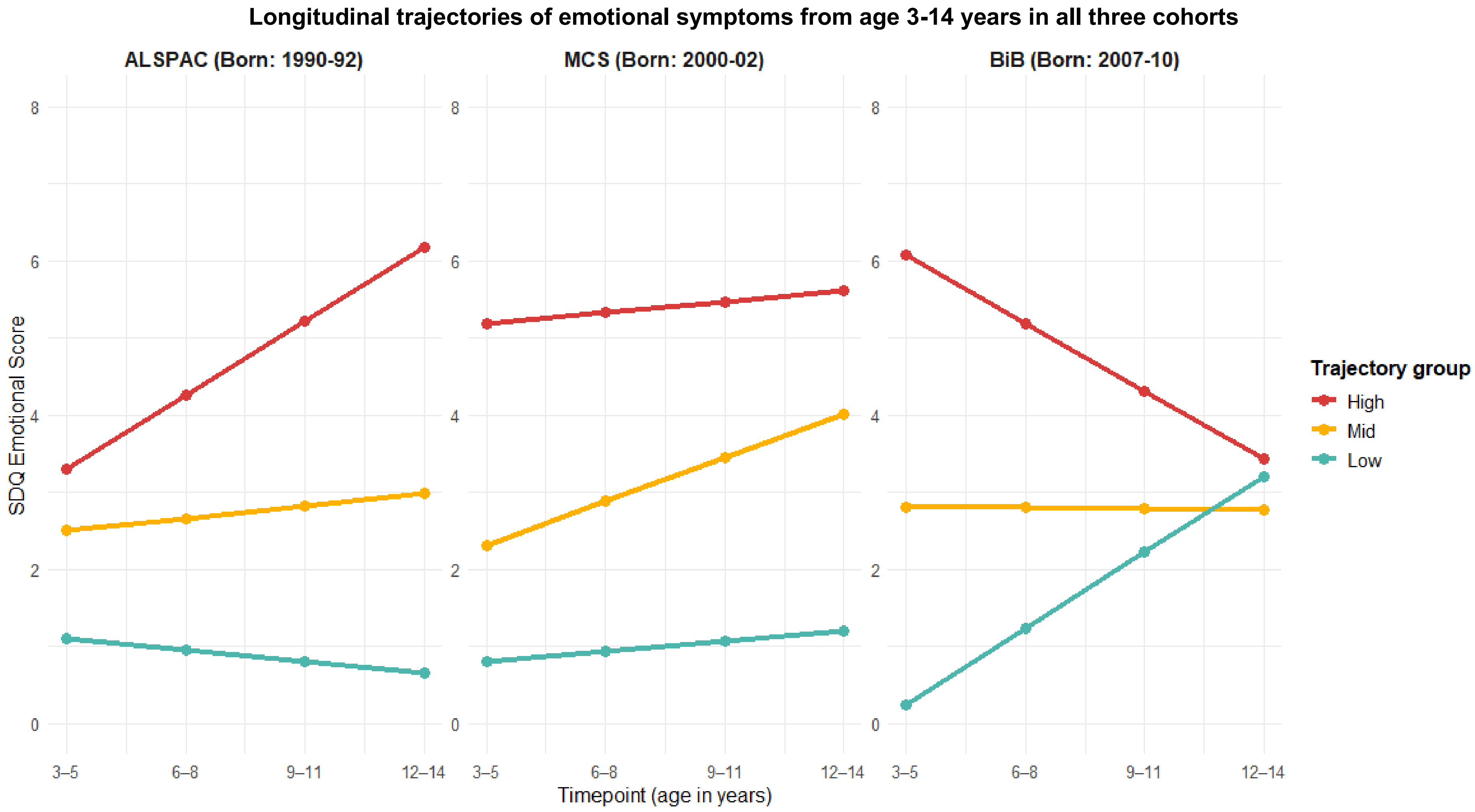

**Figure S3b. Comparison of observed and estimated means in 3 class linear models**

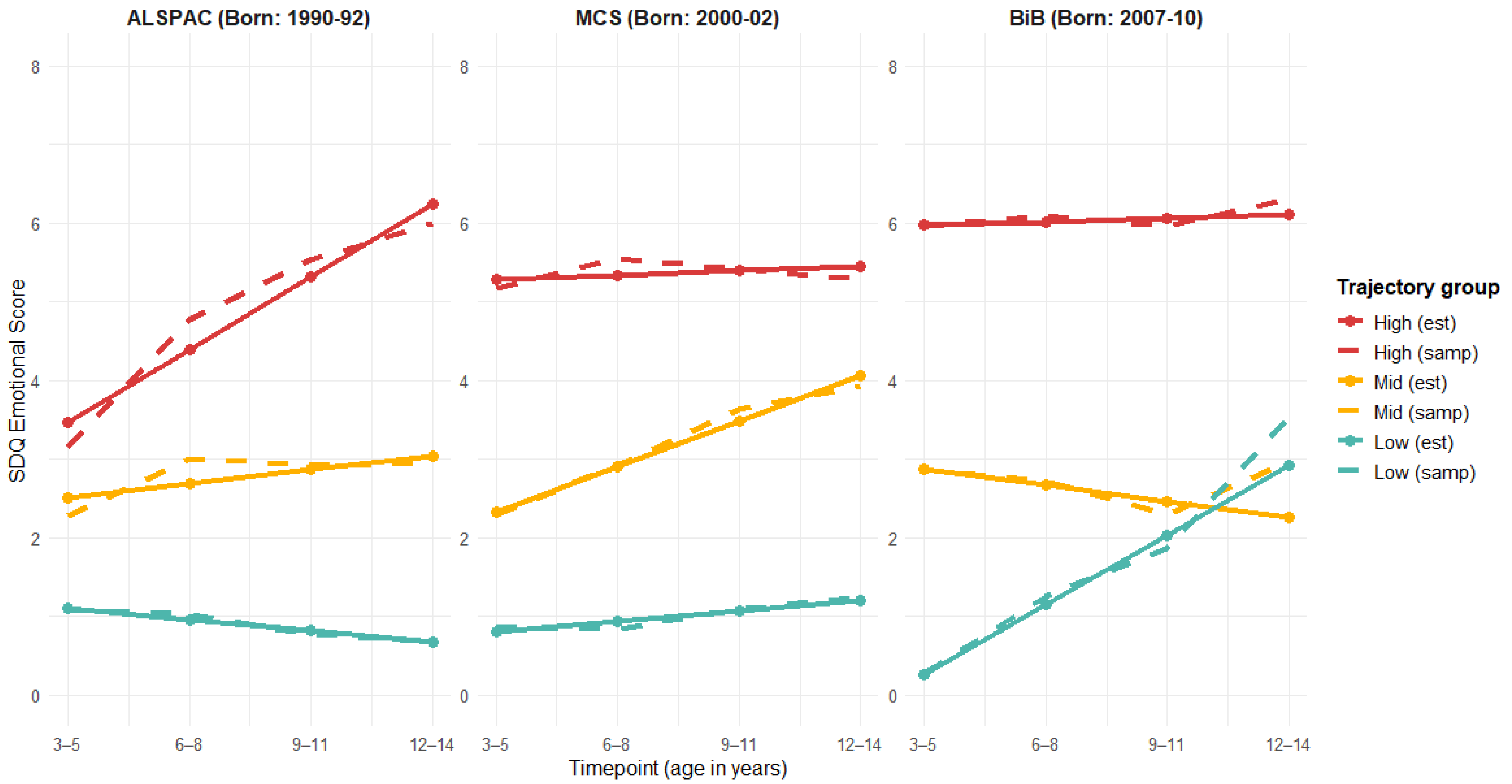

### GRoLTS-Checklist

| | Checklist Item | Reported? | Detail |
|---|---|---|---|
| **1** | Is the metric of time used in the statistical model reported? | Yes | Age in years, groupings described |
| **2** | Is information presented about the mean and variance of time within a wave? | Yes | Supplementary table S1a |
| **3a.** | Is the missing data mechanism reported? | Yes | FIML assumes MAR |
| **3b.** | Is a description provided of what variables are related to attrition/missing data? | Yes | Table S2a and S2b |
| **3c.** | Is a description provided of how missing data in the analyses were dealt with? | Yes | Methods - FIML |
| **4** | Is information about the distribution of the observed variables included? | Yes | Table 1 |
| **5** | Is the software mentioned? | Yes | Methods section |
| **6a.** | Are alternative specifications of within-class heterogeneity considered (e.g., LGCA vs. LGMM) and clearly documented? If not, was sufficient justification provided as to eliminate certain specifications from consideration? | Yes, considered and justification for LCGA reported | Justified in methods |
| **6b.** | Are alternative specifications of the between-class differences in variance/covariance matrix structure considered and clearly documented? If not, was sufficient justification provided as to eliminate certain specifications from consideration? | Yes, and justification for LCGA reported | Justified in methods |
| **7** | Are alternative shape/functional forms of the trajectories described? | Yes, Quadratic considered but justification for linear described. | Justified in methods. |
| **8** | If covariates have been used, can analyses still be replicated? | Yes | Sufficient detail provided for replication |
| **9** | Is information reported about the number of random start values and final iterations included? | Yes | Yes – main text and supplement. |
| **10** | Are the model comparison (and selection) tools described from a statistical perspective? | Yes | Yes - Supplementary tables S3-5 |
| **11** | Are the total number of fitted models reported, including a one-class solution? | Yes | Yes - Supplementary tables S3-5 |
| **12** | Are the number of cases per class reported for each model (absolute sample size, or proportion)? | Yes | Yes - Supplementary tables S3-5 |
| **13** | If classification of cases in a trajectory is the goal, is entropy reported? | Yes | Yes - Supplementary tables S3-5 |
| **14a.** | Is a plot included with the estimated mean trajectories of the final solution? | Yes | Yes - Figure 1 |
| **14b.** | Are plots included with the estimated mean trajectories for each model? | Partially | Either a plot or fit statistics given for all models. Plot for 3class model selected. Plots not given for 1,2,4,5,6 class models. |
| **14c.** | Is a plot included of the combination of estimated means of the final model and the observed individual trajectories split out for each latent class? | Yes | Figure S2b |
| **15** | Are characteristics of the final class solution numerically described (i.e., means, SD/SE, n, CI, etc.)? | Yes | Table S15. |
| **16** | Are the syntax files available (either in the appendix, supplementary materials, or from the authors)? | Partially | Will be made available on OSF or github repository following publication. |
